# Extreme temperatures and mortality in 326 Latin American cities

**DOI:** 10.1101/2021.11.16.21266420

**Authors:** Josiah L. Kephart, Brisa N. Sánchez, Jeffrey Moore, Leah H. Schinasi, Maryia Bakhtsiyarava, Yang Ju, Nelson Gouveia, Waleska T Caiaffa, Iryna Dronova, Saravanan Arunachalam, Ana V. Diez Roux, Daniel A. Rodríguez

## Abstract

**Background:** Climate change and urbanization are rapidly increasing human exposure to extreme ambient temperatures, yet few studies have examined the impact of temperature on mortality across Latin America, where 80% of residents live in urban areas.

**Methods:** We used distributed lag nonlinear conditional Poisson models to estimate city-specific associations between daily temperatures above (“heat”) and below (“cold”) each city-specific minimum mortality temperature and all-cause mortality, overall and stratified by age and cause of death. We estimated the percentage of total deaths attributable to heat or cold (excess death fraction [EDF]) and the difference in mortality per 1°C higher daily mean temperature above the 95^th^ percentile of observed daily temperature.

**Results:** We analyzed data from 326 cities in nine Latin American countries between 2002-2015, representing 15,431,532 deaths from 249 million residents. The EDF of total deaths from heat was 0.67% (95% confidence interval [CI] 0.58%, 0.74%), and from cold was 5.09% (95% CI 4.64%, 5.47%). EDFs from heat and cold were particularly high among individuals aged 65+ years (0.81% [95% CI 0.75%, 0.86%] and 6.82% [95% CI 6.41%, 7.18%], respectively). The relative risk of death per 1°C increase above the city-specific 95^th^ percentile daily temperature was 1.057 (95% CI 1.046, 1.067).

**Conclusions:** In Latin American cities, a substantial proportion of deaths are attributable to non-optimal ambient temperatures. Older populations are particularly vulnerable. Marginal increases in observed hot temperatures are associated with steep increases in mortality risk. Projected increases in temperature from climate change may have a substantial impact on mortality.

**SIGNIFICANCE STATEMENT:** Latin America has a large population at risk of urban heat exposure, yet little is known about the linkages between ambient temperature and health in the region. We analyzed over 15 million deaths in 326 Latin American cities to characterize the relationship between ambient temperature and mortality, overall and by age and cause of death. We found that 5.75% of all deaths are associated with non-optimal temperatures, older individuals are particularly vulnerable, and cardiorespiratory deaths are especially affected. A single degree increase (1°C) in daily temperature was associated with a 5.7% higher mortality among hot days, suggesting that projected increases in temperature from climate change may have a substantial impact on mortality.

## 1. Introduction

Anthropogenic greenhouse gas emissions continue to accelerate the pace of global climate change, with eight of the nine hottest years between 1880 and 2019 occurring since 2010 (1). The process of urbanization has also contributed to an increase in human exposure to extreme heat (2) in particular, through the urban heat island effect (3–5). Ambient temperatures in urban cores, where residents concentrate, can far exceed temperatures in peri-urban areas, causing urban residents to be especially exposed to extreme heat (2).

Exposure to extreme hot and cold ambient temperatures has been linked to excess morbidity and premature mortality through a range of physiological mechanisms (6). Recent global analyses (7–9) and multiple regional studies in North America (10, 11), Europe (12), and the Western Pacific (13) have reported substantial impacts of non-optimal temperatures on mortality, observing notable variations in the temperature-mortality relationship between and within world regions (9). These regional differences are likely driven by variations in the urban heat island effect (5), climate, geography, built environment, social structures, and existing adaptive capacity. Accordingly, region-specific analyses are critically needed to understand the most vulnerable subpopulations and to inform regional and local policies, emergency response plans, and climate adaptation efforts. The vast majority of regional analyses on temperature and mortality have focused on high-income countries or included only a small number of cities in the Global South (14). This reflects a paucity of research on climate and health in low- and middle-income countries more generally (15), which hampers our ability to protect health in areas with the greatest susceptibility to climate change.

Latin America is one of the most urbanized regions of the world (16) and therefore has a large population at risk of urban heat exposure (2). A few studies in Latin America have examined the relation between temperature and mortality within a single (17–20) or small number of cities (21), but examinations of the impact of ambient temperatures on health on a multinational or regional level within Latin America are lacking. A rare exception is a recent global study by Zhao et al. that included data from 66 locations in Latin America and the Caribbean, with a majority of observed cities located in two countries (Brazil and Peru) (7). In the coming decades, Latin America is projected to experience a substantial increase in mean annual temperature (22) and critically, an astounding increase in the frequency of extreme heat events (23). Between the late 20^th^ century and mid-21^st^ century, the frequency of extremely hot days (defined by 95^th^ percentile daily mean temperature between 1961-1990) in South America’s largest cities is projected to increase by five to ten times under a mid-level climate scenario (RCP4.5) (23).

Current and future public health threats from these climatic changes are exacerbated by Latin America’s rapidly ageing (24) and exceptionally urbanized population (16). These intertwined challenges of increasing population exposure (urbanization), population susceptibility (ageing population), and a warming climate make extreme temperatures a critical 21^st^ century environmental health challenge.

To better understand the relationship between temperature and mortality and to inform current and future efforts to prevent temperature-related deaths in Latin America, this study characterized the impact of non-optimal temperatures on all-cause and cause-specific mortality across 326 cities in Latin America, encompassing 98% of all cities of 100,000 persons or more in nine countries and over 15 million deaths from a total of 249 million residents.

## 2. Results

### 2.1. City characteristics

Overall and country-stratified characteristics of study cities are presented in **Table 1**. The median population among all cities was approximately 267,000 residents and 10% of cities had populations greater than 1.2 million. The proportion of the population aged 65 or more years varied between countries, with a median of 5.3% aged 65+ years among Peruvian cities and a median of 8.8% aged 65+ years among cities in Chile. The median city had 1,454 deaths per year during the study period (10^th^ percentile 715 deaths, 90^th^ percentile 6,825 deaths), and there was a grand total of 15,431,532 deaths included in the analysis. City names, average annual deaths, and temperature summaries (median, 5^th^ and 95^th^ percentile) for each city are provided as Supplemental Material (**Table S1**). The locations and annual mean temperatures of all cities are presented in **Figure 1**. In certain areas of Peru, Mexico, and elsewhere, large differences in mean temperatures are apparent in cities that are relatively close together geographically. These variations are concentrated in mountainous regions with widely varying altitude which drives these temperature differences.

**Table 1.** Population, mortality, and temperature characteristics of 326 Latin American cities

| Countries | Number of Cities | Study Period | City Population (thousands)<sup>§</sup> | Proportion Age ≥ 65<sup>§</sup> | Annual Deaths<sup>§</sup> | Mean Temperature<sup>§</sup> |
| --- | --- | --- | --- | --- | --- | --- |
| All countries | 326 | 2002-2015 | 267 (130, 1,292) | 6.6 (4.6, 9.1) | 1,454 (715, 6,825) | 21.3 (14.9, 25.9) |
| Argentina | 28 | 2009-2015 | 334 (133, 1,398) | 8.2 (7.0, 12.0) | 2,234 (821, 12,028) | 17.5 (14.4, 21.6) |
| Brazil | 152 | 2002-2015 | 221 (121, 1,292) | 7.0 (4.8, 8.8) | 1,443 (717, 6,279) | 22.2 (18.9, 26.4) |
| Central America* | 10 | 2009-2015 | 264 (185, 2,773) | 7.2 (4.7, 8.4) | 1,639 (1,085, 14,416) | 23.8 (14.4, 25.8) |
| Chile | 21 | 2004-2015 | 210 (140, 920) | 8.8 (7.0, 10.7) | 1,088 (771, 5,215) | 13.7 (10.8, 17.0) |
| Mexico | 92 | 2005-2015 | 352 (142, 1,089) | 5.6 (4.4, 7.0) | 1,825 (775, 5,272) | 20.3 (15.6, 25.8) |
| Peru | 23 | 2008-2015 | 288 (131, 885) | 5.3 (3.9, 6.8) | 978 (442, 3,644) | 19.6 (8.0, 24.7) |

§ Median (10th, 90th Percentiles)
\*The Central America group in this analysis consists of cities in Guatemala, Panama, Costa Rica, and El Salvador

**Figure 1.**
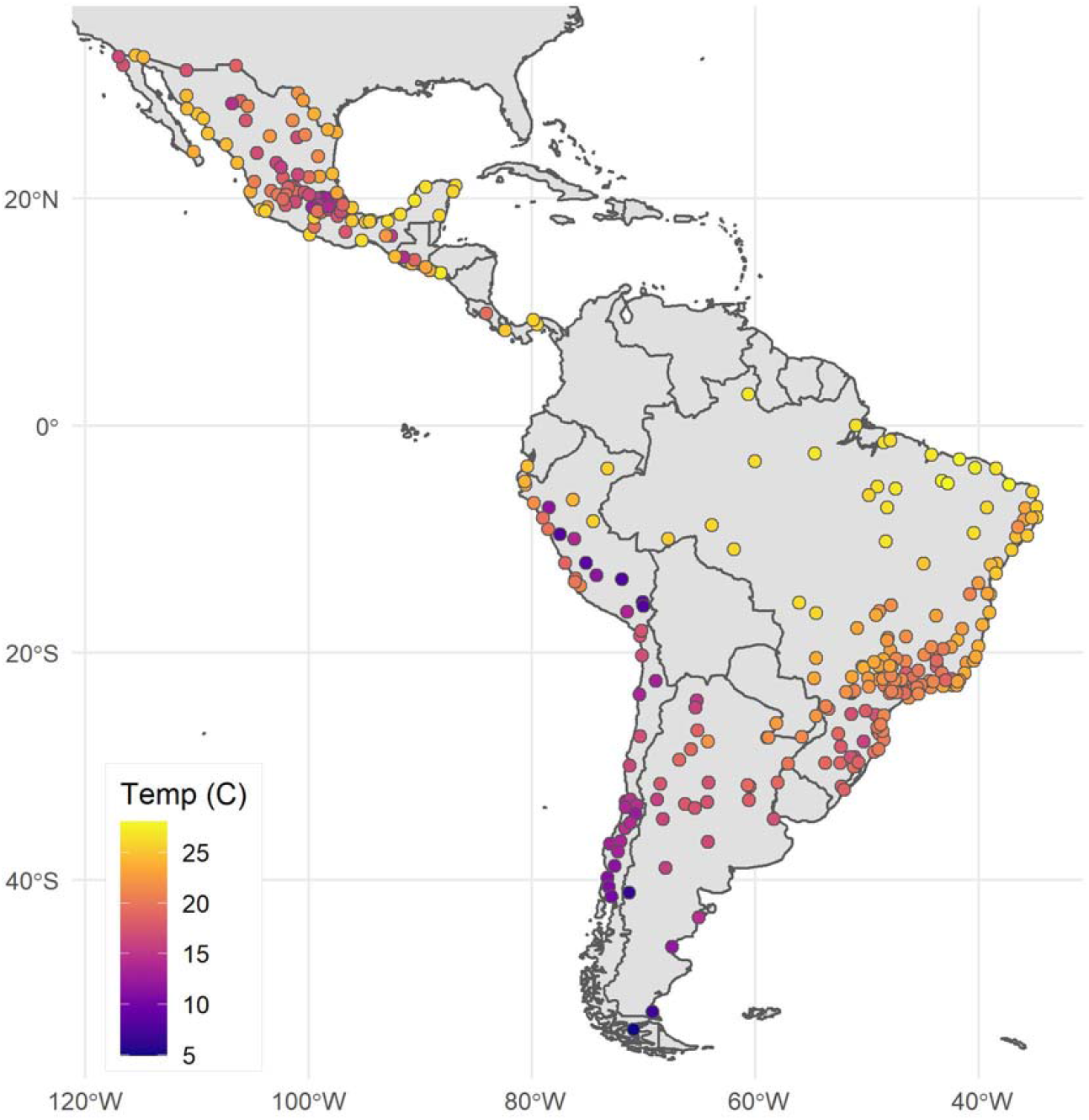
Annual mean temperatures during city-specific observation period in 326 Latin American cities

### 2.2. Association between temperature and all-cause mortality

**Figure 2** shows estimated city-specific temperature-mortality associations alongside histograms of the daily temperature distribution for six cities selected to illustrate between-city differences in the temperature distributions and in the shape of the temperature-mortality associations. Similar plots for all cities are presented in the Supplementary Material (**Figure S1)**. Most cities had an approximate U-shaped relationship between temperature and mortality, although the relation between temperature and mortality was not symmetric on both sides of the optimal temperature. At temperatures below the optimal temperature mortality increased gradually as temperatures dropped, while at temperatures above the optimal temperature mortality increased more steeply as temperatures rose. The sharp increase in mortality with increasing temperatures was most pronounced for cities that regularly exceed approximately 25°C (e.g. Buenos Aires, Mérida, and Rio de Janeiro in **Figure 2**). However, among cities with temperate or cold climates that rarely (or never) exceeded approximately 25°C (e.g. Lima, Mexico City, and Los Angeles in **Figure 2**) mortality did not increase or increased only minimally as temperatures increased.

The overall relative risk of all-cause mortality (all ages) per 1°C increase in daily mean temperature for temperatures above the city specific 95^th^ percentile was 1.057 (95% confidence interval [CI] 1.046 to 1.067). However, this estimate varied geographically (**Figure 3)**. We observed a high proportion of cities with large increases in mortality per 1°C higher temperature under extreme heat (relative risk [RR] >=1.050, red and purple dots) in relatively temperate areas of southern Brazil, Argentina, and parts of Mexico. We found a high proportion of cities with minimal changes in mortality under city-specific extreme heat (RR<=1.000, yellow dots) among cities in the high-altitude Andes (relatively cold) and in northeast Brazil (tropical with relatively low temperature variability).

**Figure 2.**
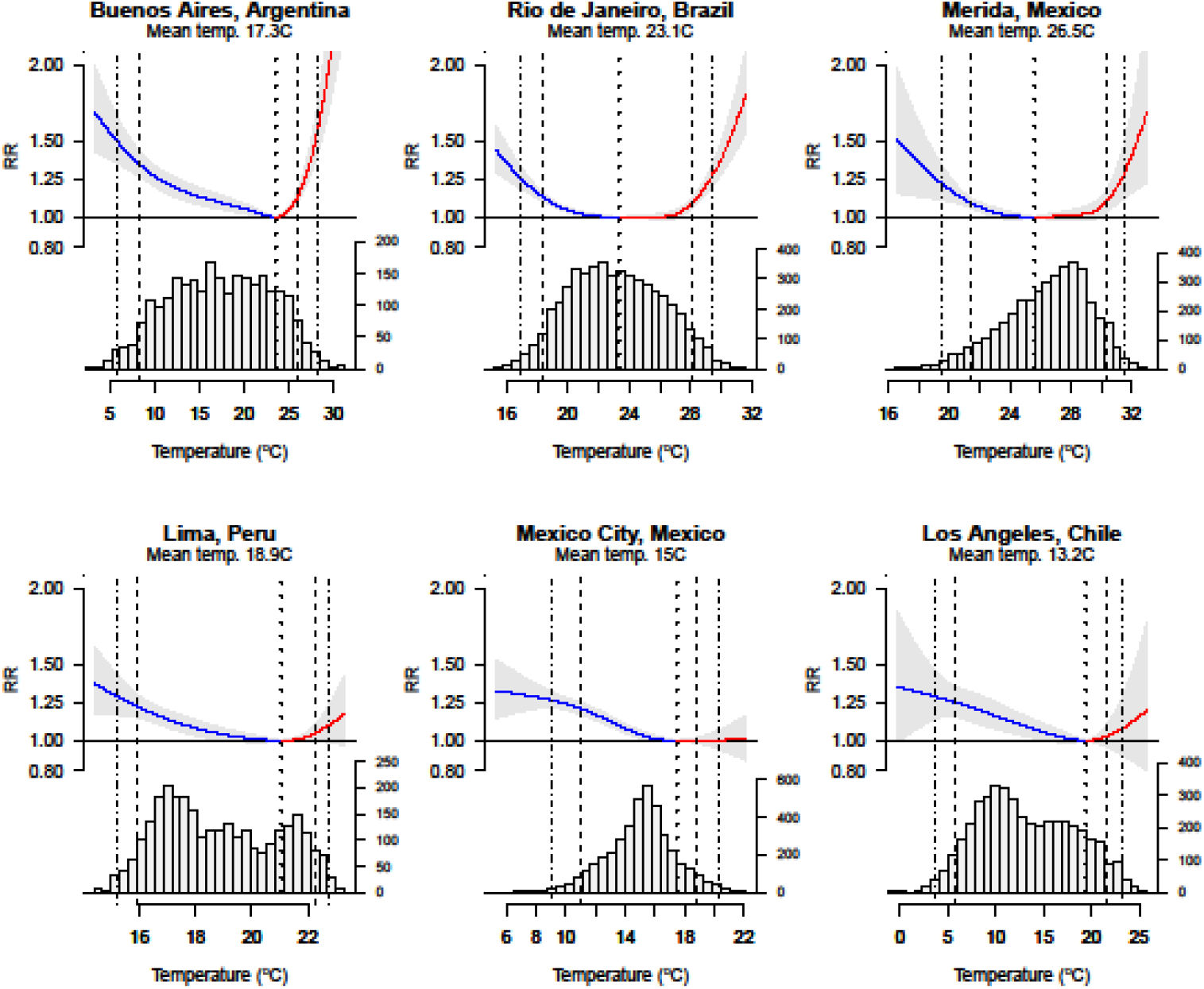
The city-specific temperature-mortality exposure-response association (accumulated over 21 days) and distribution of daily temperatures for six selected cities. Vertical lines are placed at the optimal (i.e. minimum mortality) temperature (dotted), the 5^th^ and 95^th^ percentiles of the temperature distribution (dashed), and 1^st^ and 99^th^ temperature percentiles (dash-dot). Blue parts of the curves correspond to the city-specific temperature-mortality associations for temperatures below the minimum mortality temperature (cold); red curves denote temperature-mortality associations for temperatures above the minimum mortality temperature (heat).

**Figure 3.**
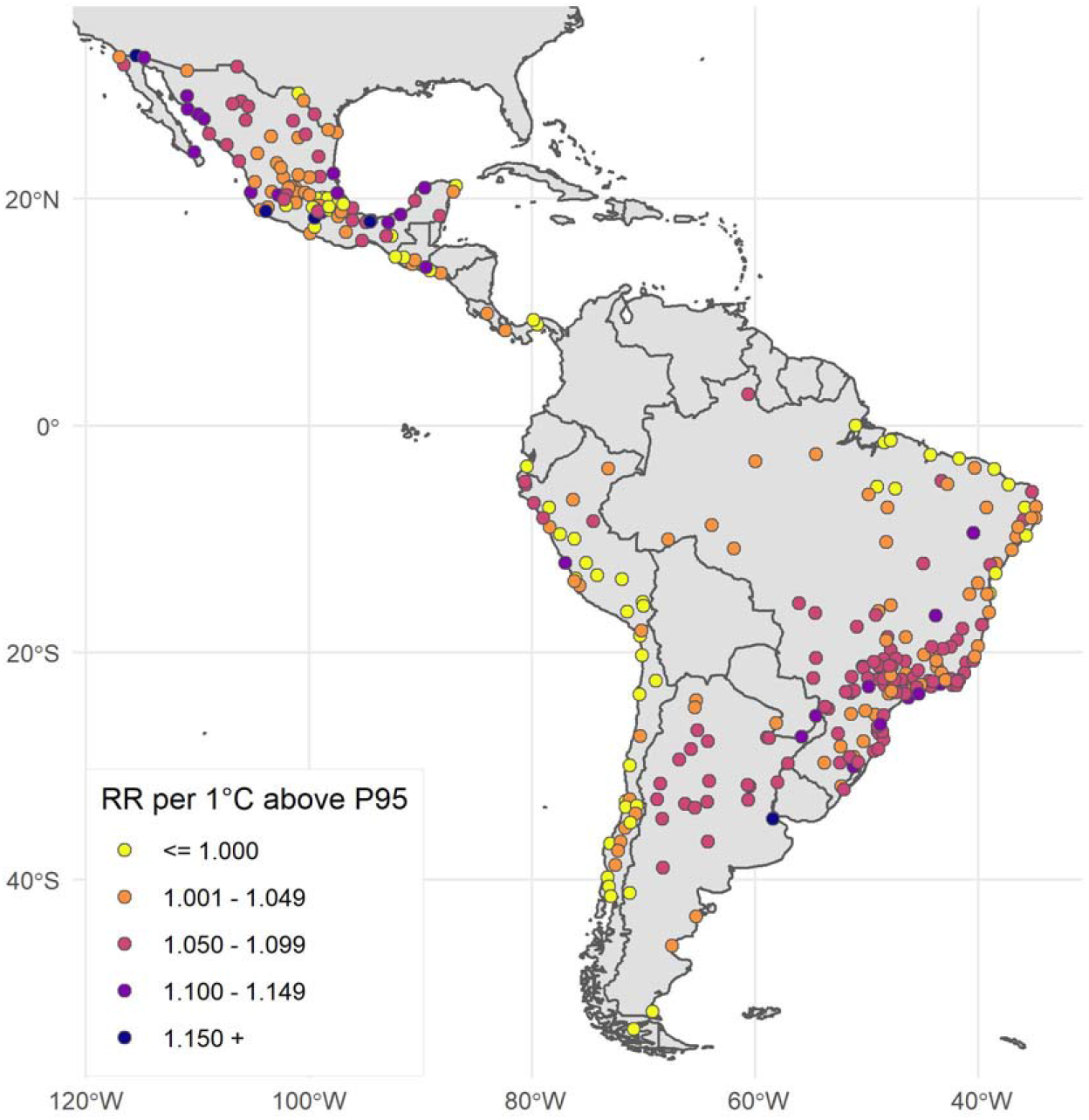
City-specific relative risk (RR) of heat-related mortality per 1°C increase above the 95^th^ percentile observed daily temperature in 326 Latin American cities

The relative risk of mortality per 1°C decrease in extreme cold temperatures (< 5^th^ percentile) was 1.034 (95% CI 1.028 to 1.040). A map of the city-specific relative risk per 1°C decrease during extreme cold (temperature below the 5^th^ percentile) is presented in the Supplemental Material (**Figure S2**).

Estimates of excess deaths from non-optimal temperatures, overall and by age and cause of death groupings, are presented in **Table 2**. We estimate that 5.75% (95% CI 5.31 to 6.07) of deaths at all ages from all causes are associated with non-optimal temperatures. The excess death fraction (EDF) for heat (the cumulative effect of all temperatures above optimal) was 0.67% (95% CI 0.58 to 0.74), with extreme heat (≥95^th^ percentile city-specific observed temperatures) contributing a substantial portion of the heat-related excess deaths (0.42%, 95% CI 0.38 to 0.45). The EDF from cold was substantially higher than heat, at 5.09% (95% CI 4.66 to 5.42) for all cold and 1.03% (95% CI 0.99 to 1.06) for extreme cold (≤5^th^ percentile city-specific observed temperatures). The EDF from non-optimal temperatures was consistently higher among individuals aged 65 + than among the total population (**Table 2**). City-specific minimum mortality temperature and EDFs for cold and heat among all-ages are presented in the Supplementary Material in **Table S1**.

**Table 2.**
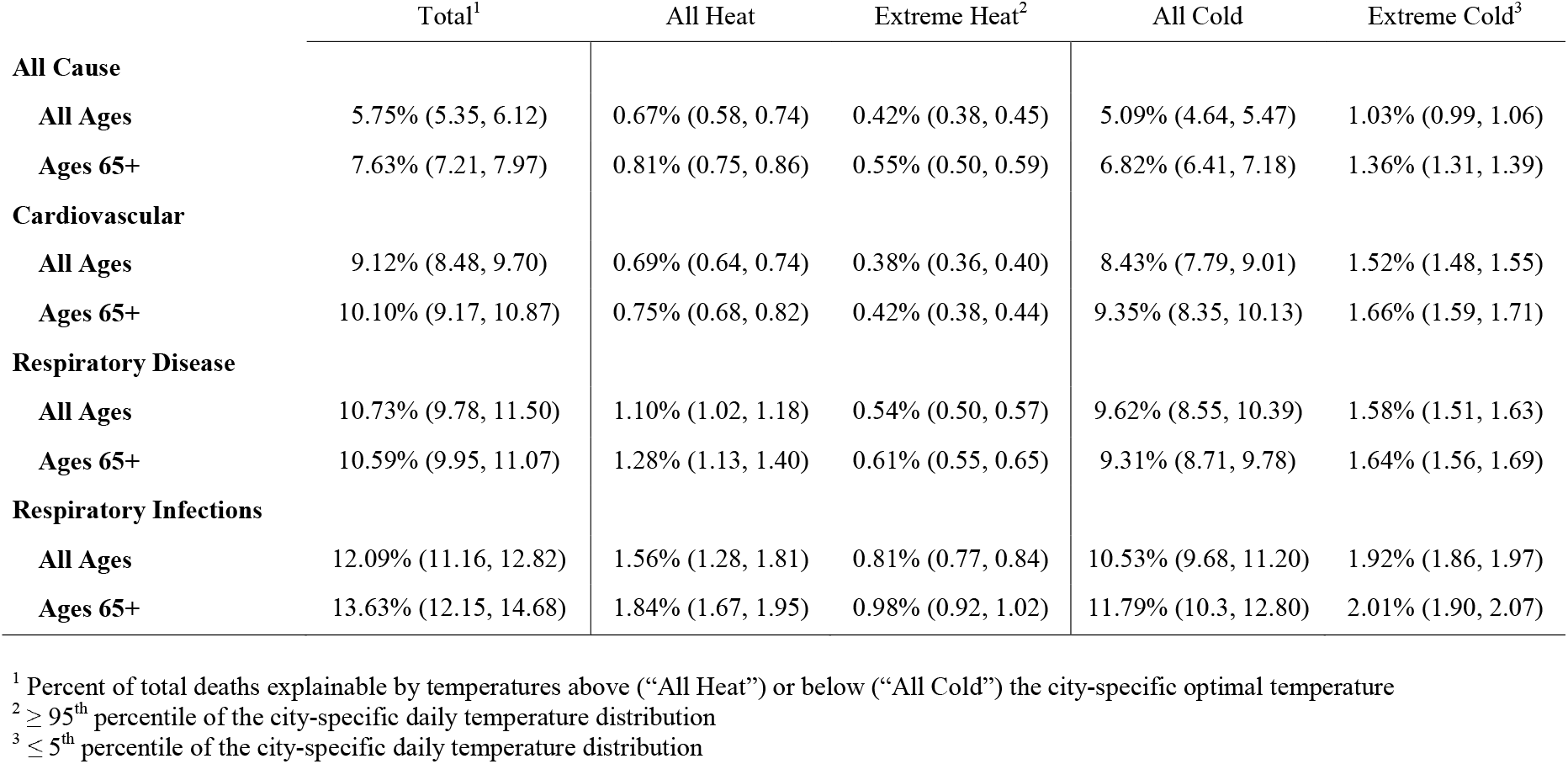
Excess death fraction associated with non-optimal temperatures with 95% confidence intervals

### 2.3. Associations between temperature and cause-specific mortality

Compared to deaths from all causes and all ages, we observed a substantially higher EDF from non-optimal temperatures for cardiovascular disease (9.12% [95% CI 8.48, 9.70]), respiratory diseases (10.73% [9.78, 11.50]), and respiratory infections (12.09% [11.16, 12.82]) **(Table 2)**. The same pattern (higher EDFs for cardiovascular and respiratory deaths than for all-cause mortality) was observed for cold temperatures. In the case of hot temperatures, EDFs for respiratory mortality were slightly higher than for all-cause mortality, but EDFs for cardiovascular mortality were similar to those observed for all-cause mortality. Patterns were similar for deaths at all ages and over 65 years, although the EDFs for over 65 were slightly higher than those for all ages.

The cause of death most strongly associated with both heat and cold was respiratory infections, with 1.56% (95% CI 1.28 to 1.81) of all-ages deaths attributable to heat and 10.53% (95% CI 9.68 to 11.20) attributable to cold.

## 3. Discussion

We examined the contribution of ambient temperature to age- and cause-specific mortality across 326 cities in highly urbanized Latin America between 2002 - 2015. We found that a substantial proportion of mortality was attributable to ambient cold and to a lesser extent heat. This mortality burden is larger among older individuals and among deaths from cardiorespiratory causes. Importantly, we found that even small increases in extreme heat can rapidly increase mortality risk.

Overall, a substantially higher proportion of deaths is attributable to ambient cold than to ambient heat, which corroborates findings from similar analyses in other settings. A recent analysis by Zhao et al. estimated temperature-mortality associations in 750 locations from 43 countries (including 66 locations in Latin American and the Caribbean), and extrapolated these estimates globally at 0.5° x 0.5° grid size (approximately 55km x 55km at the equator) using meta-predictors (7). This study reported global EDFs of 8.52% for cold and 0.91% for heat for all-age all-cause mortality. Within Latin American and the Caribbean only, this same study estimated EDFs of 4.71% from cold and 1.06% from heat. This is similar to our estimated EDFs of 5.09% from cold and 0.67% from heat from observations in 326 Latin American cities.

Drawing conclusions regarding the relative impact of heat and cold on mortality risk using EDFs is rendered complex by the fact that the EDF estimate is driven by both the impact of temperature changes on mortality as well as by the distribution of hot and cold days. Even though mortality risk increases more gradually with decreasing temperatures below the optimal temperature than with increasing temperatures above the optimal temperature, the EDF attributable to cold is larger because there are generally more days below than above the optimal temperature. For this reason, we complemented the well-established methods for estimating temperature-related EDFs by calculating the difference in mortality risk per 1°C increase under extreme temperatures above the city-specific 95^th^ percentile (extreme heat) and per 1°C decrease below the 5^th^ percentile (extreme cold), representing the hottest and coldest 18 days within a typical year at each setting. Although mortality risk increased in a dose-response fashion both below and above optimal temperatures, the increase per 1°C difference was substantially larger for extreme heat than for extreme cold temperatures (5.7% vs. 3.4% respectively). These findings suggest that shifting temperature distributions to higher levels may at least initially result in pronounced increases in mortality risk as extreme heat becomes more frequent.

We also observed that the increase in mortality associated with a 1°C increase in extremely hot temperatures has substantial geographic variation. For example, increases in mortality risk per 1°C increase in extreme heat are particularly steep in the cities of coastal Mexico, northern Argentina, and southern Brazil. Residents of these areas may be particularly vulnerable to extreme heat now and in the near-term under even marginal increases in the frequency of extreme heat from climate change. A greater understanding of the city level factors (physical social or policy characteristics) that explain this heterogeneity may help identify effective actions to buffer future impacts of temperature increases.

Our results support the prevailing understanding that individuals aged 65+ years are more vulnerable to heat-related mortality (19, 25) and further emphasize that older individuals require prioritization within efforts to protect the public from extreme ambient temperatures. Compared to all-cause mortality, we found higher proportions of temperature-related deaths caused by cardiovascular diseases, non-communicable respiratory diseases, and respiratory infections. The proportion of deaths attributable to both heat and cold were higher for respiratory deaths than for all deaths combined. We found similar heat-related EDFs for cardiovascular deaths (0.69% [95% CI 0.64 to 0.74]) and deaths from all causes (0.67% [95% CI 0.58 to 0.74), counter to other findings that cardiovascular mortality is a specific risk from heat exposure (26). Further analyses with more refined measures of cause of death may shed light on the reasons for these results.

A key strength of this study is the inclusion of all cities of 100,000 or more residents in nine countries of Latin America. We used two-phase statistical methods which allow us to capture local specificities while also permitting the pooling of information across multiple cities to derive more valid and reliable estimates of the impact of heat on mortality. An important byproduct of our approach is the generation of city-specific temperature mortality curves, as well as the extension of this analysis to summarize the city-specific and overall changes in risks with marginal increases of extreme temperatures. This study also used population-weighted daily temperature reanalysis, which provides more spatially resolved estimates of true population exposure compared to the more common approach of applying measurements from one or a few temperature monitors to an entire city population (27). Furthermore, we used individual-level mortality data, allowing us to examine variation in mortality by age and cause of death within a large number of cities in the Global South. Like other work, our exposure measure is limited as it does not account for inter-individual differences in exposures. Physiological exposure to heat varies widely between individuals by access to green vegetation, access to air-conditioning or other cooling mechanisms, characteristics of urban form, and time-activity-location patterns (28). Another limitation of our analysis is that we were not able to control for time-varying confounders, such as air pollution, although for pollutants whose levels are affected by temperature, air pollution may be a mediator rather than a confounder. In addition, there is evidence of two-way effect modification of the health impacts of air pollution and temperature that may influence our results (29–31). These interactions could have differential impacts on our estimates of heat-versus cold-related mortality due to seasonal variations in concentrations of different air pollutants, which may also vary by location (32).

Within a large and diverse sample of cities in the most urbanized region of the Global South, a substantial proportion of deaths can be attributed to ambient temperatures. These temperature-related deaths are particularly concentrated in older populations and are more common for deaths from cardiorespiratory diseases. Moreover, we observed that marginal increases in context-specific hot temperatures dramatically increase the risk of mortality. While during the study period (2002 – 2015) cold temperatures contribute to more deaths than warm temperatures, our analysis affirms that the projected climate-related increases in the frequency of extremely hot days would likely substantially increase the risk of heat-related deaths across the region.

Although precise temperature modelling estimates are needed to derive quantitative estimates of expected changes in deaths linked to global warming under different scenarios, our results suggest that rising temperatures will increase mortality above optimal temperatures by shifting more days to temperatures above the optimal temperature generally but especially by shifting more days to levels above the 95^th^ percentile (extreme heat), where heat-related deaths increase rapidly with increases in temperature. Policymakers in Latin America and elsewhere must prioritize actions to prevent present and future health risks of extreme temperatures.

## 4. Methods

### 4.1. Study area

This study was conducted as part of the *Salud Urbana en América Latina* (SALURBAL) project. The SALURBAL project has compiled and harmonized data on environmental, social, and heath characteristics for all cities of 100,000 residents or more (a total of 371 cities) in 11 Latin American countries (33). Cities in SALURBAL were defined as urban agglomerations with more than 100,000 residents in 2010 (34), allowing for examination of a range of city sizes from small cities to megacities. These cities are composed of clusters of administrative units encompassing the visually apparent urban built up area as identified using satellite imagery (34). In this analysis, we include 326 cities in Argentina, Brazil, Chile, Costa Rica, El Salvador, Guatemala, Mexico, Panama, and Peru. Cities in Colombia and Nicaragua were excluded due to limited availability of mortality data.

### 4.2. Data sources

We used the ERA5-Land climate reanalysis with ∼9km horizontal resolution (35) to estimate the population-weighted daily mean ambient temperature for each city from 2002 – 2015. We calculated daily mean temperatures by averaging ERA5-Land hourly temperatures by calendar days. Because ERA5-Land omits grid cells that contain >50% water, 99 of 326 cities (30%) contained ≥1 grid cells with missing temperature predictions within the city boundaries (mean % missing grid cells among 99 cities with ≥1 missing pixels: 18%). We imputed temperature values for missing grid cells using a random forest regression model that included resampled ERA5 temperature (36) (31 km resolution), elevation, and aspect (compass direction that terrain faces), with further modeling of the residuals using kriging spatial interpolation. To better approximate population exposures, we spatially weighted city temperature using 2010 estimates of the spatial distribution of the population (WorldPop, https://www.worldpop.org/: Argentina, Brazil, Chile, Costa Rica, El Salvador, Guatemala, and Mexico) or urban footprint (Global Urban Footprint, https://www.un-spider.org/node/11424: Panama and Peru).

Individual-level mortality data were compiled from vital registration systems in each country. Mortality records included date of death, municipality of residence, age at death, and cause of death using the International Classification of Diseases v.10 (37). We applied World Health Organization Global Health Estimate classifications (38) to categorize deaths into the following groupings: all-cause (GHE tiers I., II., III.), cardiovascular (II.G.), non-communicable respiratory disease (II.H.), and respiratory infections (I.B.). We stratified deaths by age at death (<65 and 65+ years).

We compiled city-level population characteristics including total population and population age composition from Census Bureaus, National Institutes of Statistics, or similar sources for each country. More information on this data compilation has been previously published (34).

### 4.3. Statistical analysis

We estimated the temperature-mortality associations for deaths at all ages and stratified by age at death (<65 and 65+ years).

We modeled the nonlinear relationship between daily mean temperature and all-cause mortality using a two-stage approach (9, 39). First, for each city, we constructed a time series of daily all-cause and cause-specific mortality counts by aggregating individual-level mortality records within each city and age group (<65 and 65+ years), and linked it to the corresponding population-weighted daily mean temperature. We used distributed lag (0-21 days) nonlinear conditional Poisson models to estimate city-specific, non-linear associations between mortality and daily temperature (39, 40). The non-linear associations were estimated using natural cubic splines with knots placed at the minimum, maximum, and 10^th^, 75^th^, and 90^th^ percentiles of the city-specific distribution of daily temperatures. We selected this number of knots and their locations based on their wide use in prior literature (9), and because they yielded equivalent or better model fit compared to others. The models conditioned on strata defined by day of the week, month and year, offering strong control for seasonality and secular changes, and yielding inferences based on short-term temperature variability. Second, we combined city-specific estimates using a random effects meta-regression to obtain smoothed (numerically stabilized) non-linear association curves, given that some smaller cities have less precision due to their relatively few deaths. The dependent variables in the meta-regression were the four reduced spline coefficients which can be used to reconstruct the temperature-mortality association curve summed across lags (41); the meta-predictors were each city’s median observed daily temperature, temperature range, and country (Central American cities were treated as a single group due to the small number of study cities in each country).

The smoothed curves were used to estimate each city’s ‘optimal’ temperature, defined as the observed temperature where the temperature-mortality association curve achieved its minimum value (9). The smoothed curves were displayed graphically for each city, and, to communicate risk numerically, used to obtain two complementary types of summaries. First, we estimated excess death fractions (EDF) from non-optimal temperatures compared to the optimal temperature (also known as attributable fraction). The EDF was defined as the ratio of estimated temperature-related excess deaths to total deaths for each city throughout the study period, expressed as a percent. We estimated the EDF attributable to temperatures above or below the city-specific optimal temperature, as well as the EDF associated with either extreme heat or cold, defined as ≥95^th^ or ≤5^th^ percentiles of city-specific daily temperatures, respectively. Second, for each city, we approximated the steepness or ‘slope’ of the nonlinear association curve under extreme heat (≥95^th^ percentile temperature) and extreme cold (≤5^th^ percentile), expressed as relative risk per one Celsius degree (°C) higher temperature. To obtain this ‘slope’ for extreme heat, we extracted the log-relative risk of mortality at the 99^th^ compared to the 95^th^ percentile of the city-specific observed distribution of daily temperatures and divided it by the difference in Celsius degrees between the 99^th^ and 95^th^ percentile of the temperature distribution. We also estimated ‘slopes’ for extreme cold temperatures by dividing the log-relative risk at the 1^st^ compared to the 5^th^ percentiles of the city’s temperature distribution by the difference in °C between the 1^st^ and 5^th^ percentiles. While the EDF has the advantage of combining information about the relative risk and the observed number of days above or below the optimal temperature, the second type of summary is useful in communicating risk in more intuitive units (i.e., difference in risk per degree higher temperature).

We also estimated temperature-mortality associations with deaths from cause-specific groupings: cardiovascular disease, non-communicable respiratory disease, and respiratory infections. Due to low counts of daily cause-specific deaths for many smaller cities, instead of city-specific analyses we created 12 groups of cities with similar temperature distributions and estimated nonlinear temperature-mortality associations for each group. Cities were grouped via hierarchical clustering using Ward’s minimum variance method, with each city’s cumulative temperature distribution serving as inputs. Compared to clustering only on a handful of temperature summaries (e.g., city’s temperature mean and variance), this approach groups cities based on the full temperature distribution. The same conditional Poisson models were used, with the addition of city identifier when forming strata to condition on (i.e., strata formed by city ID, year, month and day of the week). A map of the city cluster groupings used in the cause-specific analysis is presented in the Supplemental Material (**Figure S3**).

All analyses were performed in R (version 3.6.0) and modelling was done with the mvmeta (41), dnlm (42), and gnm (43) packages. Clustering of cities was performed using the factoextra package (44).

## Data Availability

Vital registration and population data for Brazil, Chile, and Mexico were downloaded from publicly available repositories from statistical agencies in each country. Vital registration and population data for Argentina, Costa Rica, El Salvador, Guatemala, Panama and Peru were obtained directly from statistical agencies in each country. A link to these agency websites can be accessed via https://drexel.edu/lac/data-evidence/data-acknowledgements/.

## Acknowledgements and funding sources

This study was financially supported by the Wellcome Trust [216029/Z/19/Z], [205177/Z/16/Z]. The authors thank Xavier Delclòs-Alió, PhD for his contributions to the study design and coordination. The authors acknowledge the contribution of all SALURBAL project team members. For more information on SALURBAL and to see a full list of investigators, see https://drexel.edu/lac/salurbal/team/. SALURBAL acknowledges the contributions of many different agencies in generating, processing, facilitating access to data or assisting with other aspects of the project. Please visit https://drexel.edu/lac/data-evidence for a complete list of data sources. The funding sources had no role in the analysis, writing, or decision to submit the manuscript.

## Author contributions

DAR and AVDR conceptualized the analysis, acquired funding, and provided supervision. YJ, SA, DAR, and AVDR led Data Curation. BNS and JM completed the formal analysis. JLK created the original draft of the manuscript. All authors contributed to methodology and manuscript editing.

## Data availability

Requests for the code used in this study should be made to the corresponding author.

