## Supplemental Information for "Extreme temperatures and mortality in 326 Latin American cities"

**This PDF file includes:**

Figures S1 to S3
Table S1

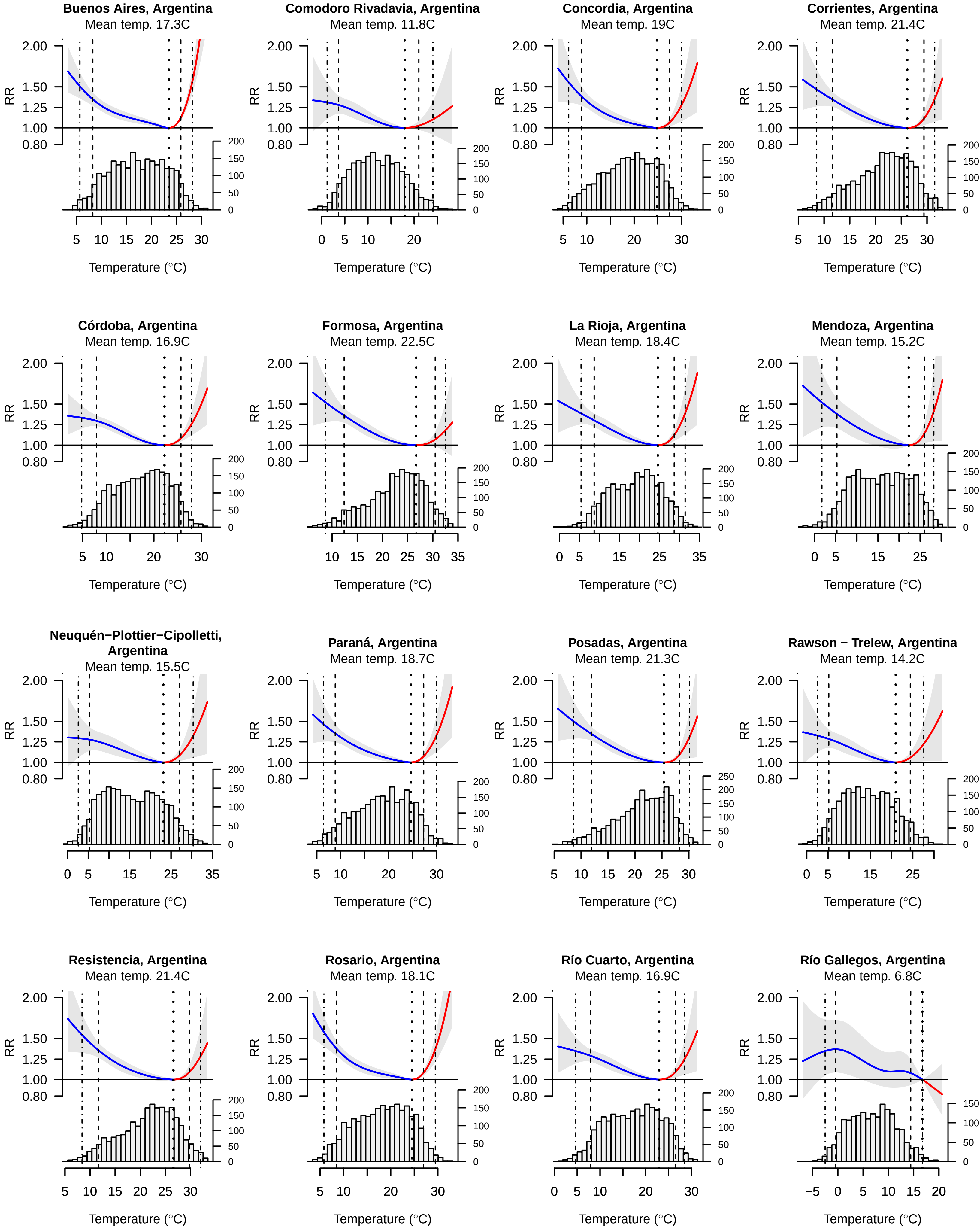
Buenos Aires, Argentina
Mean temp. 17.3C
Comodoro Rivadavia, Argentina
Mean temp. 11.8C
Concordia, Argentina
Mean temp. 19C
Corrientes, Argentina
Mean temp. 21.4C
Córdoba, Argentina
Mean temp. 16.9C
Formosa, Argentina
Mean temp. 22.5C
La Rioja, Argentina
Mean temp. 18.4C
Mendoza, Argentina
Mean temp. 15.2C
Neuquén−Plottier−Cipolletti, Argentina
Mean temp. 15.5C
Paraná, Argentina
Mean temp. 18.7C
Posadas, Argentina
Mean temp. 21.3C
Rawson − Trelew, Argentina
Mean temp. 14.2C
Resistencia, Argentina
Mean temp. 21.4C
Rosario, Argentina
Mean temp. 18.1C
Río Cuarto, Argentina
Mean temp. 16.9C
Río Gallegos, Argentina
Mean temp. 6.8C
RR
Temperature (°C)

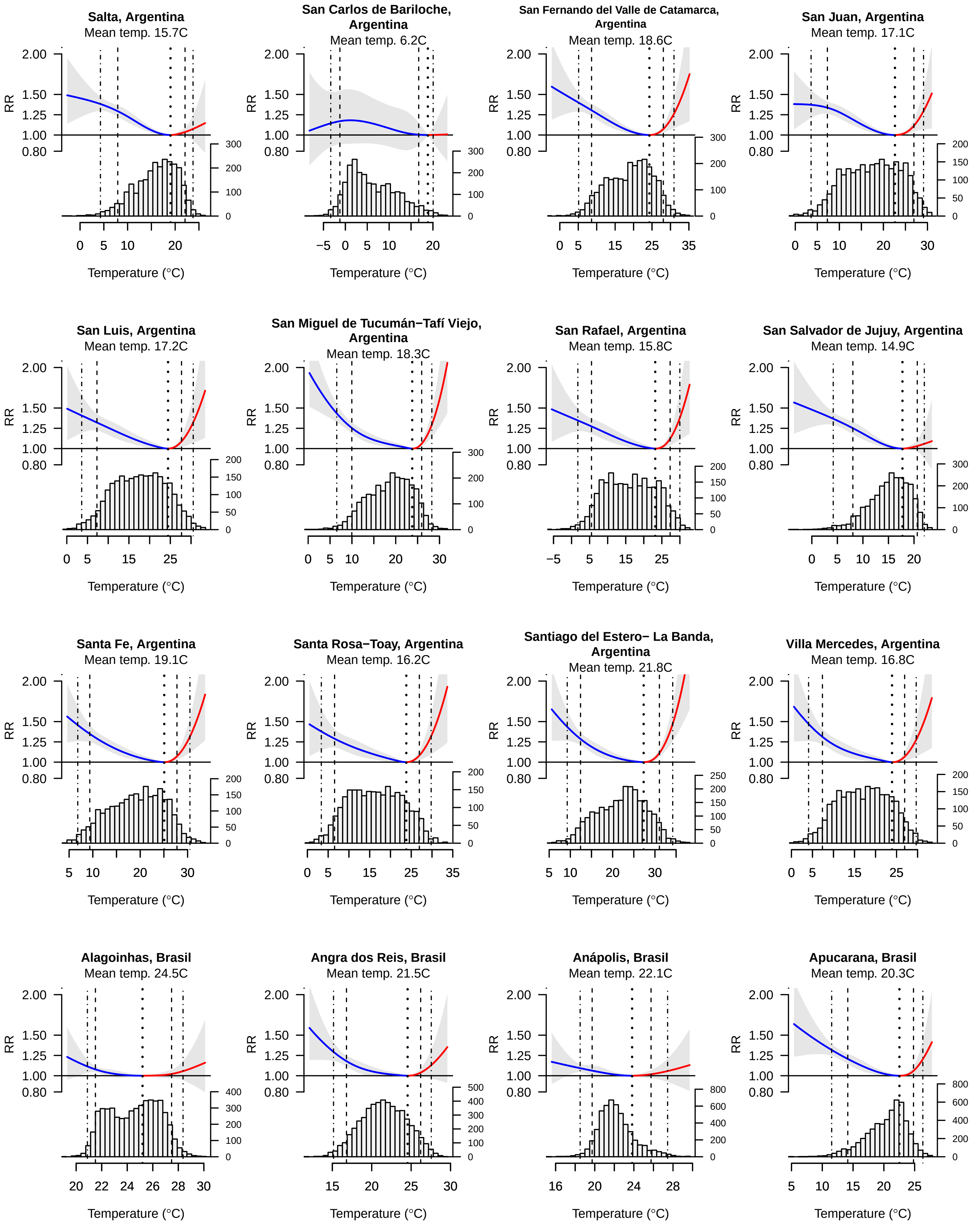
Salta, Argentina
Mean temp. 15.7C
San Carlos de Bariloche, Argentina
Mean temp. 6.2C
San Fernando del Valle de Catamarca, Argentina
Mean temp. 18.6C
San Juan, Argentina
Mean temp. 17.1C
San Luis, Argentina
Mean temp. 17.2C
San Miguel de Tucumán−Tafí Viejo, Argentina
Mean temp. 18.3C
San Rafael, Argentina
Mean temp. 15.8C
San Salvador de Jujuy, Argentina
Mean temp. 14.9C
Santa Fe, Argentina
Mean temp. 19.1C
Santa Rosa−Toay, Argentina
Mean temp. 16.2C
Santiago del Estero− La Banda, Argentina
Mean temp. 21.8C
Villa Mercedes, Argentina
Mean temp. 16.8C
Alagoinhas, Brasil
Mean temp. 24.5C
Angra dos Reis, Brasil
Mean temp. 21.5C
Anápolis, Brasil
Mean temp. 22.1C
Apucarana, Brasil
Mean temp. 20.3C
RR
Temperature (°C)

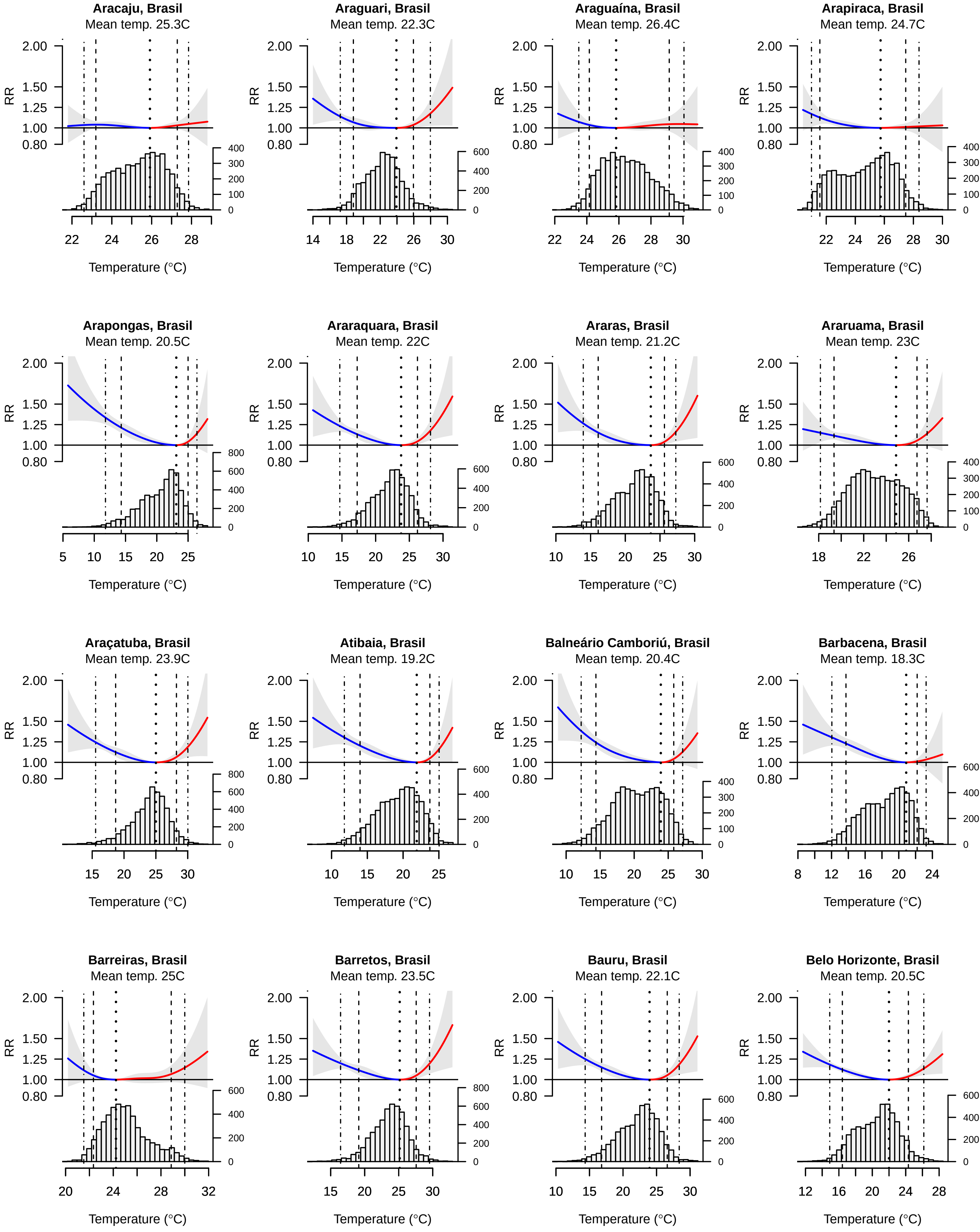
Aracaju, Brasil
Mean temp. 25.3C
Araguari, Brasil
Mean temp. 22.3C
Araguaína, Brasil
Mean temp. 26.4C
Arapiraca, Brasil
Mean temp. 24.7C
Arapongas, Brasil
Mean temp. 20.5C
Araraquara, Brasil
Mean temp. 22C
Araras, Brasil
Mean temp. 21.2C
Araruama, Brasil
Mean temp. 23C
Araçatuba, Brasil
Mean temp. 23.9C
Atibaia, Brasil
Mean temp. 19.2C
Balneário Camboriú, Brasil
Mean temp. 20.4C
Barbacena, Brasil
Mean temp. 18.3C
Barreiras, Brasil
Mean temp. 25C
Barretos, Brasil
Mean temp. 23.5C
Bauru, Brasil
Mean temp. 22.1C
Belo Horizonte, Brasil
Mean temp. 20.5C
RR
Temperature (°C)

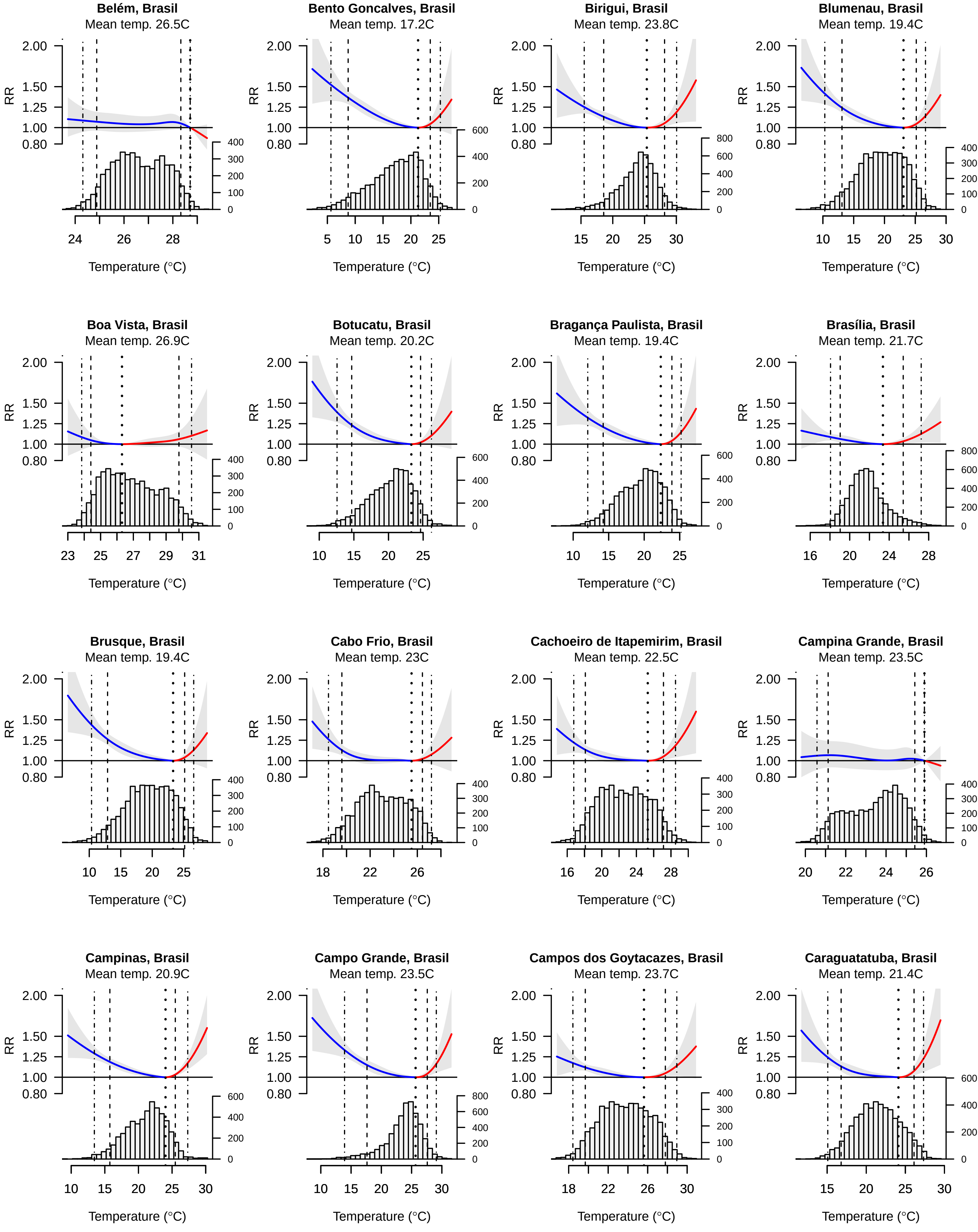
Belém, Brasil
Mean temp. 26.5C
Bento Goncalves, Brasil
Mean temp. 17.2C
Birigui, Brasil
Mean temp. 23.8C
Blumenau, Brasil
Mean temp. 19.4C
Boa Vista, Brasil
Mean temp. 26.9C
Botucatu, Brasil
Mean temp. 20.2C
Bragança Paulista, Brasil
Mean temp. 19.4C
Brasília, Brasil
Mean temp. 21.7C
Brusque, Brasil
Mean temp. 19.4C
Cabo Frio, Brasil
Mean temp. 23C
Cachoeiro de Itapemirim, Brasil
Mean temp. 22.5C
Campina Grande, Brasil
Mean temp. 23.5C
Campinas, Brasil
Mean temp. 20.9C
Campo Grande, Brasil
Mean temp. 23.5C
Campos dos Goytacazes, Brasil
Mean temp. 23.7C
Caraguatatuba, Brasil
Mean temp. 21.4C
RR
Temperature (°C)

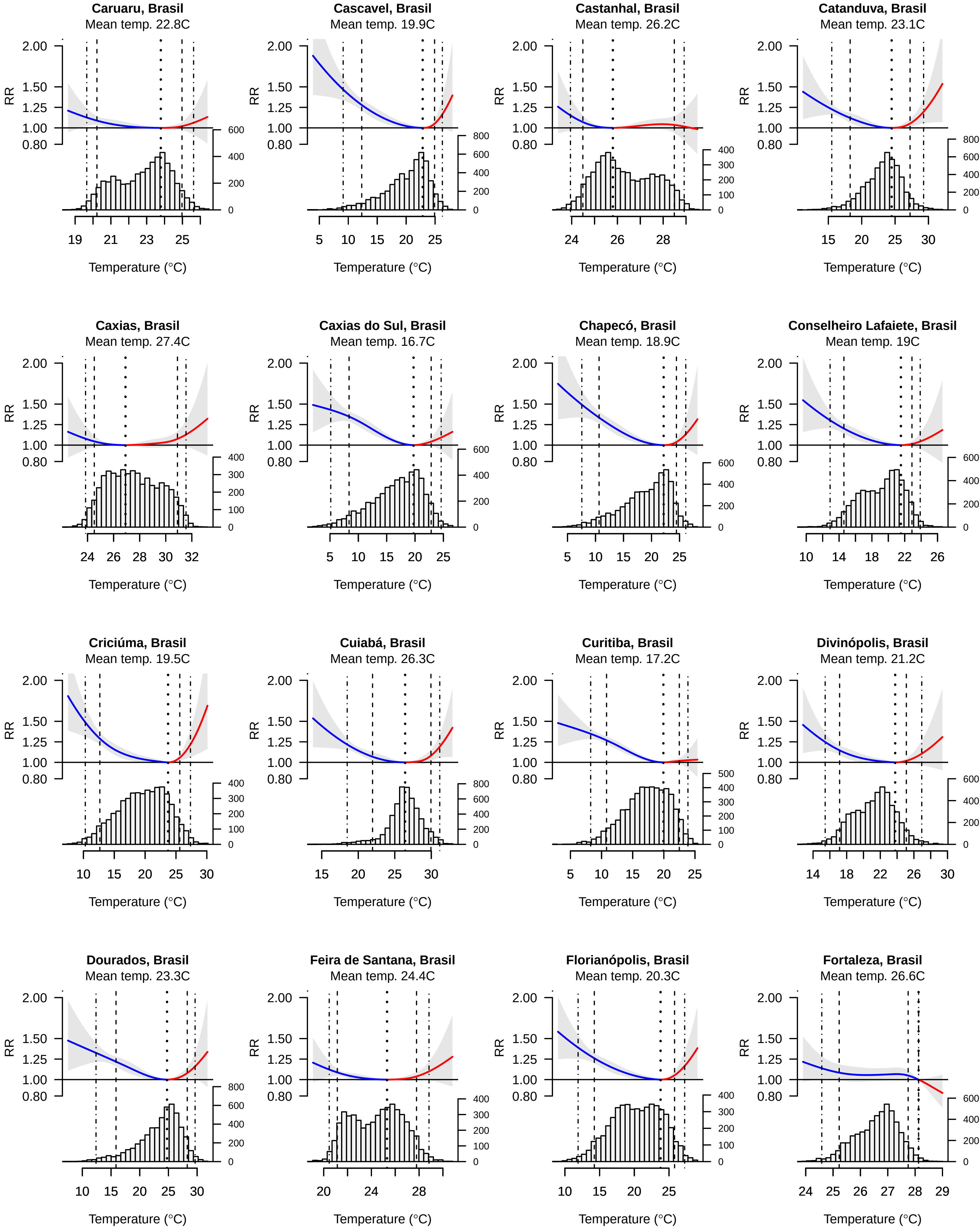

Caruaru, Brasil
Mean temp. 22.8C
Cascavel, Brasil
Mean temp. 19.9C
Castanhal, Brasil
Mean temp. 26.2C
Catanduva, Brasil
Mean temp. 23.1C
Caxias, Brasil
Mean temp. 27.4C
Caxias do Sul, Brasil
Mean temp. 16.7C
Chapecó, Brasil
Mean temp. 18.9C
Conselheiro Lafaiete, Brasil
Mean temp. 19C
Criciúma, Brasil
Mean temp. 19.5C
Cuiabá, Brasil
Mean temp. 26.3C
Curitiba, Brasil
Mean temp. 17.2C
Divinópolis, Brasil
Mean temp. 21.2C
Dourados, Brasil
Mean temp. 23.3C
Feira de Santana, Brasil
Mean temp. 24.4C
Florianópolis, Brasil
Mean temp. 20.3C
Fortaleza, Brasil
Mean temp. 26.6C
RR
Temperature (°C)

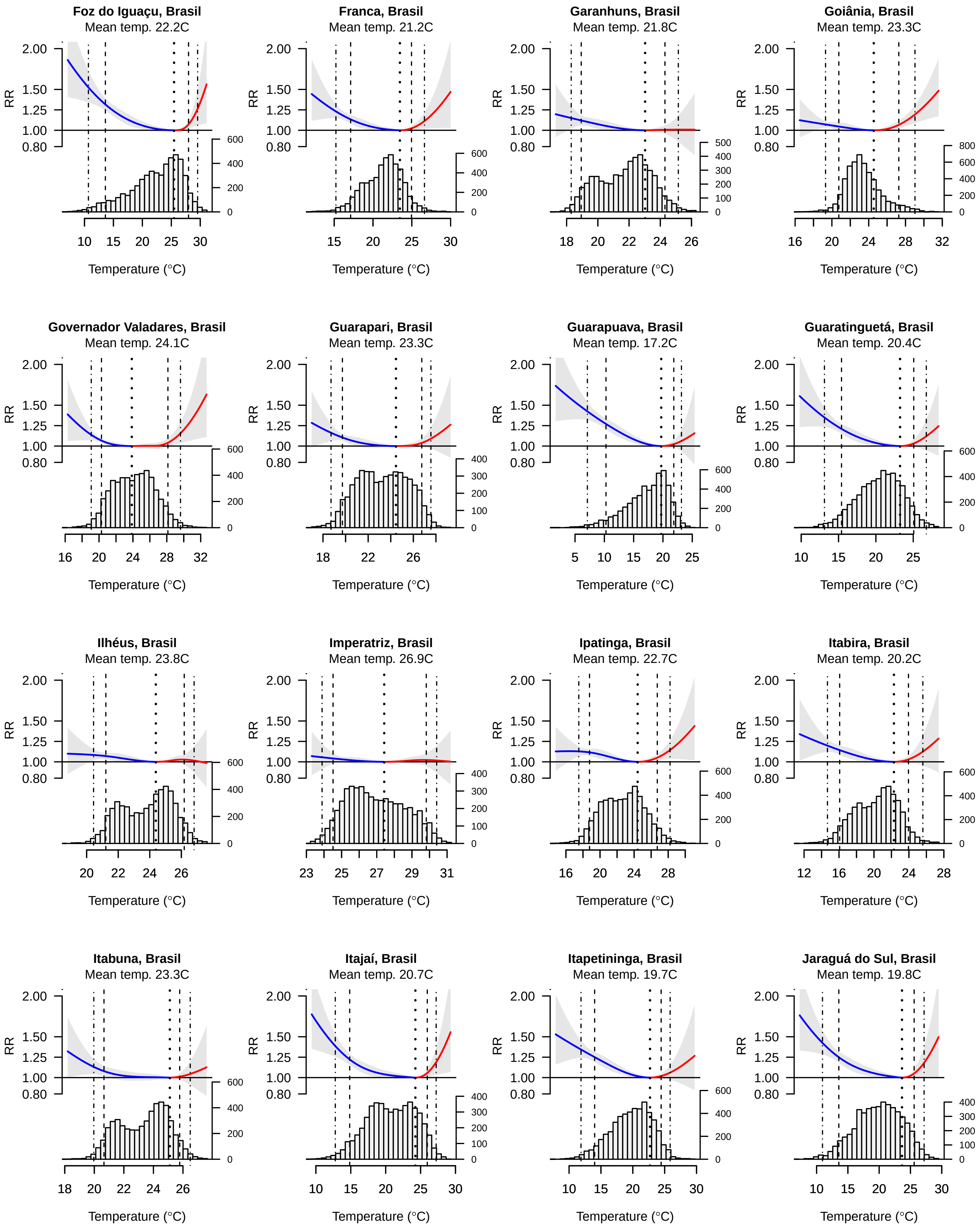

Foz do Iguaçu, Brasil
Mean temp. 22.2C
Franca, Brasil
Mean temp. 21.2C
Garanhuns, Brasil
Mean temp. 21.8C
Goiânia, Brasil
Mean temp. 23.3C
Governador Valadares, Brasil
Mean temp. 24.1C
Guarapari, Brasil
Mean temp. 23.3C
Guarapuava, Brasil
Mean temp. 17.2C
Guaratinguetá, Brasil
Mean temp. 20.4C
Ilhéus, Brasil
Mean temp. 23.8C
Imperatriz, Brasil
Mean temp. 26.9C
Ipatinga, Brasil
Mean temp. 22.7C
Itabira, Brasil
Mean temp. 20.2C
Itabuna, Brasil
Mean temp. 23.3C
Itajaí, Brasil
Mean temp. 20.7C
Itapetininga, Brasil
Mean temp. 19.7C
Jaraguá do Sul, Brasil
Mean temp. 19.8C
RR
2.00
1.50
1.25
1.00
0.80
Temperature (°C)

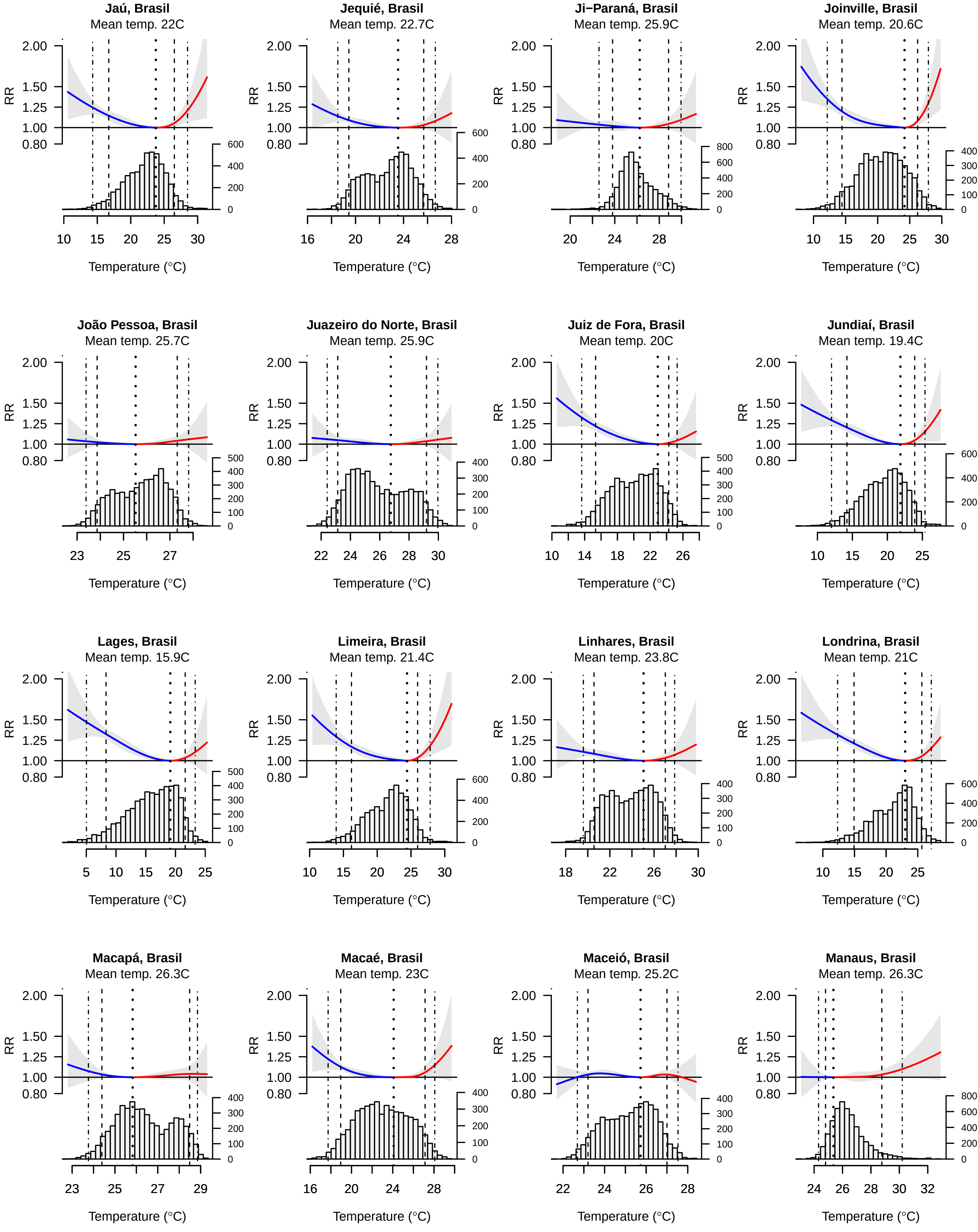
Jaú, Brasil
Mean temp. 22C
Jequié, Brasil
Mean temp. 22.7C
Ji−Paraná, Brasil
Mean temp. 25.9C
Joinville, Brasil
Mean temp. 20.6C
João Pessoa, Brasil
Mean temp. 25.7C
Juazeiro do Norte, Brasil
Mean temp. 25.9C
Juiz de Fora, Brasil
Mean temp. 20C
Jundiaí, Brasil
Mean temp. 19.4C
Lages, Brasil
Mean temp. 15.9C
Limeira, Brasil
Mean temp. 21.4C
Linhares, Brasil
Mean temp. 23.8C
Londrina, Brasil
Mean temp. 21C
Macapá, Brasil
Mean temp. 26.3C
Macaé, Brasil
Mean temp. 23C
Maceió, Brasil
Mean temp. 25.2C
Manaus, Brasil
Mean temp. 26.3C
RR
Temperature (°C)

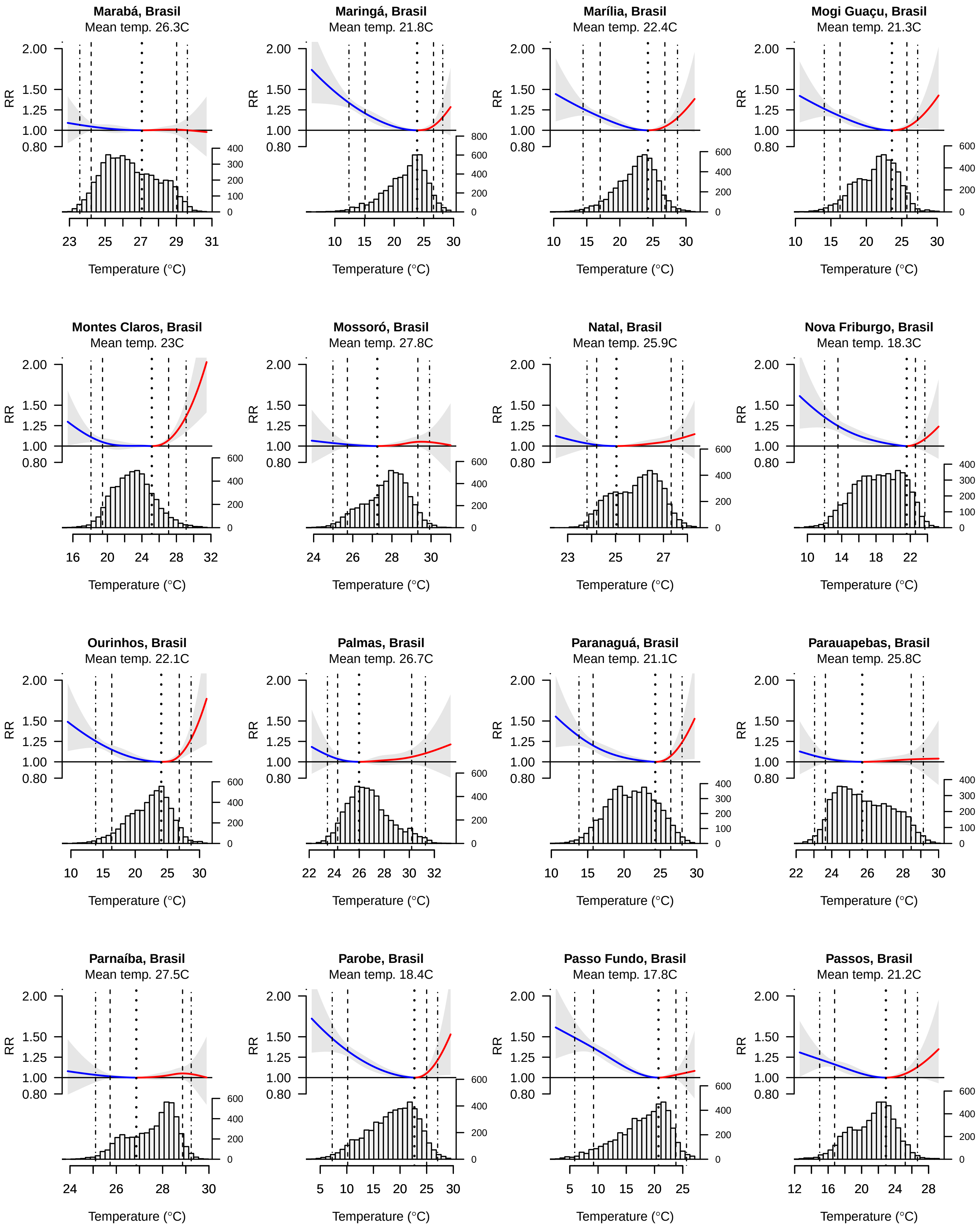
Marabá, Brasil
Mean temp. 26.3C
Maringá, Brasil
Mean temp. 21.8C
Marília, Brasil
Mean temp. 22.4C
Mogi Guaçu, Brasil
Mean temp. 21.3C
Montes Claros, Brasil
Mean temp. 23C
Mossoró, Brasil
Mean temp. 27.8C
Natal, Brasil
Mean temp. 25.9C
Nova Friburgo, Brasil
Mean temp. 18.3C
Ourinhos, Brasil
Mean temp. 22.1C
Palmas, Brasil
Mean temp. 26.7C
Paranaguá, Brasil
Mean temp. 21.1C
Parauapebas, Brasil
Mean temp. 25.8C
Parnaíba, Brasil
Mean temp. 27.5C
Parobe, Brasil
Mean temp. 18.4C
Passo Fundo, Brasil
Mean temp. 17.8C
Passos, Brasil
Mean temp. 21.2C
RR
2.00
1.50
1.25
1.00
0.80
Temperature (°C)

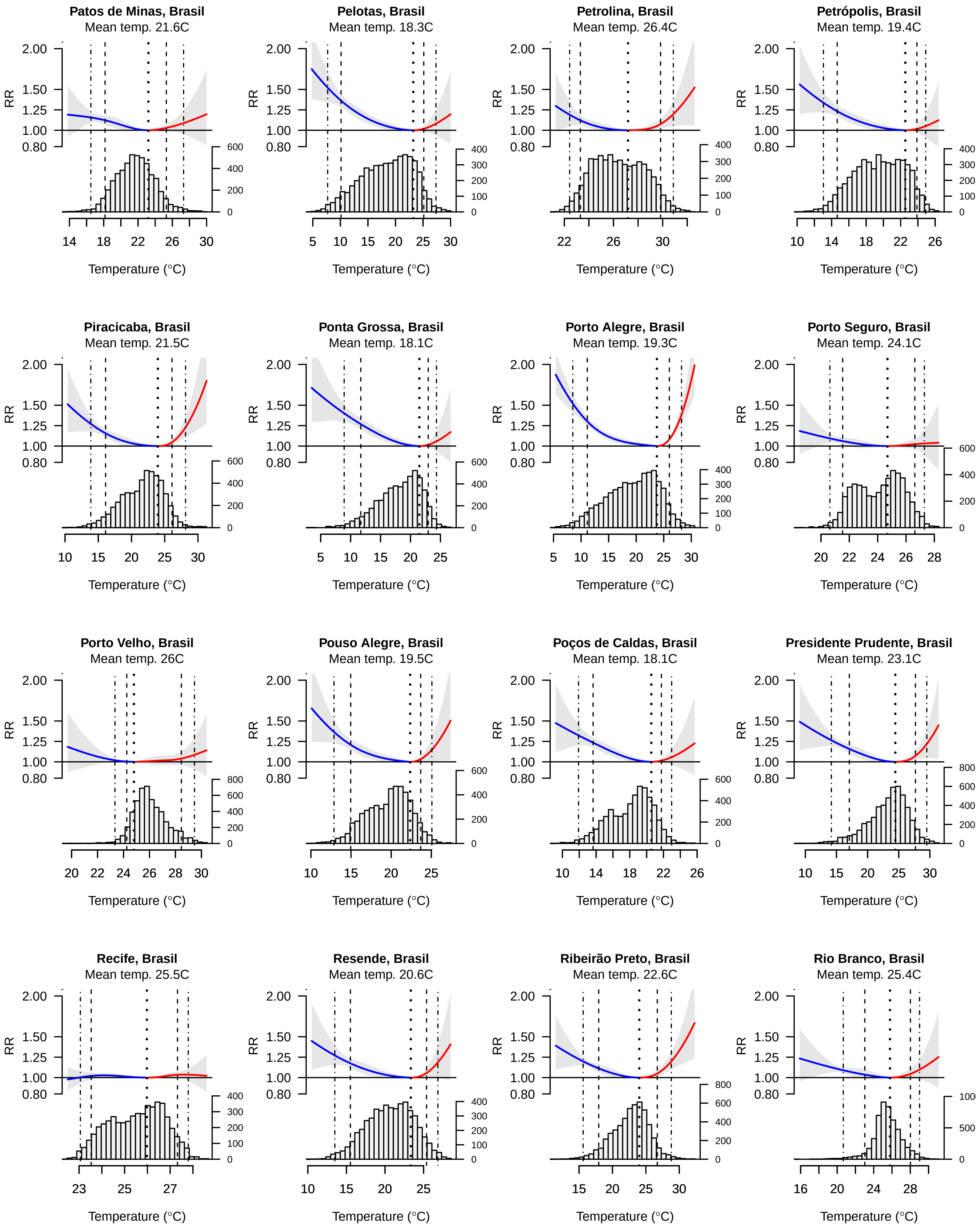
Patos de Minas, Brasil
Mean temp. 21.6C
Pelotas, Brasil
Mean temp. 18.3C
Petrolina, Brasil
Mean temp. 26.4C
Petrópolis, Brasil
Mean temp. 19.4C
Piracicaba, Brasil
Mean temp. 21.5C
Ponta Grossa, Brasil
Mean temp. 18.1C
Porto Alegre, Brasil
Mean temp. 19.3C
Porto Seguro, Brasil
Mean temp. 24.1C
Porto Velho, Brasil
Mean temp. 26C
Pouso Alegre, Brasil
Mean temp. 19.5C
Poços de Caldas, Brasil
Mean temp. 18.1C
Presidente Prudente, Brasil
Mean temp. 23.1C
Recife, Brasil
Mean temp. 25.5C
Resende, Brasil
Mean temp. 20.6C
Ribeirão Preto, Brasil
Mean temp. 22.6C
Rio Branco, Brasil
Mean temp. 25.4C
RR
2.00
1.50
1.25
1.00
0.80
Temperature (°C)

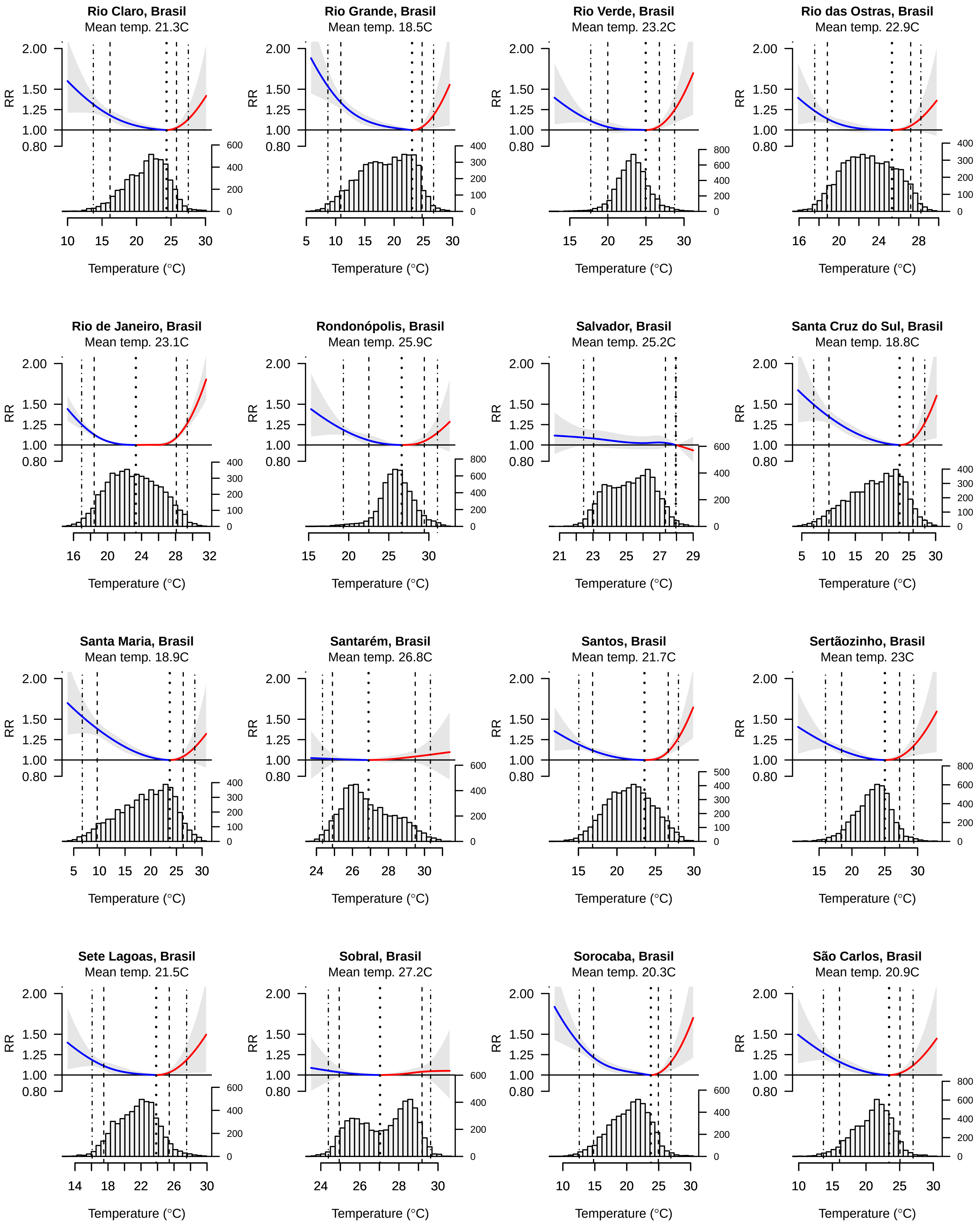
Rio Claro, Brasil
Mean temp. 21.3C
Rio Grande, Brasil
Mean temp. 18.5C
Rio Verde, Brasil
Mean temp. 23.2C
Rio das Ostras, Brasil
Mean temp. 22.9C
Rio de Janeiro, Brasil
Mean temp. 23.1C
Rondonópolis, Brasil
Mean temp. 25.9C
Salvador, Brasil
Mean temp. 25.2C
Santa Cruz do Sul, Brasil
Mean temp. 18.8C
Santa Maria, Brasil
Mean temp. 18.9C
Santarém, Brasil
Mean temp. 26.8C
Santos, Brasil
Mean temp. 21.7C
Sertãozinho, Brasil
Mean temp. 23C
Sete Lagoas, Brasil
Mean temp. 21.5C
Sobral, Brasil
Mean temp. 27.2C
Sorocaba, Brasil
Mean temp. 20.3C
São Carlos, Brasil
Mean temp. 20.9C
RR
Temperature (°C)

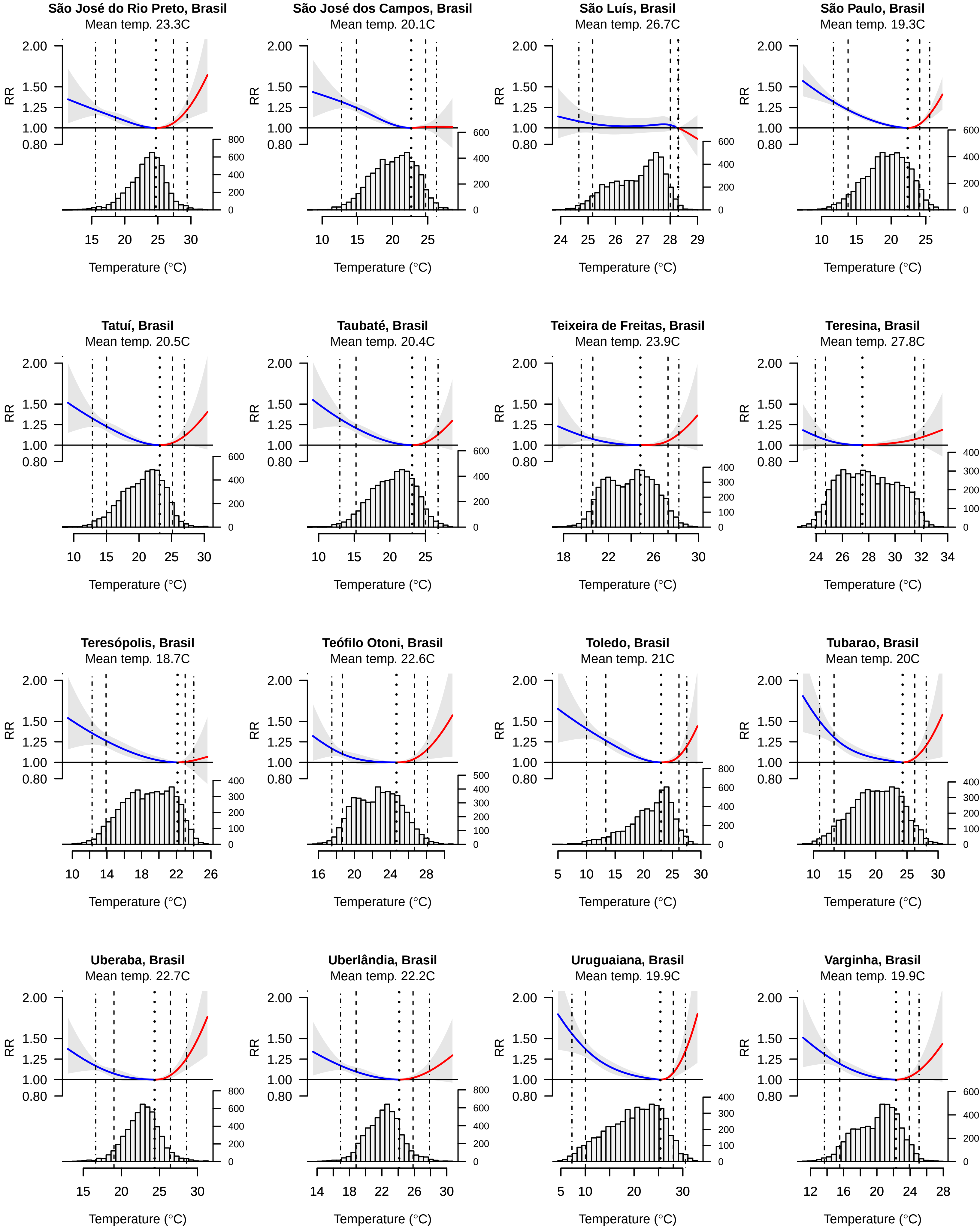
São José do Rio Preto, Brasil
Mean temp. 23.3C
São José dos Campos, Brasil
Mean temp. 20.1C
São Luís, Brasil
Mean temp. 26.7C
São Paulo, Brasil
Mean temp. 19.3C
Tatuí, Brasil
Mean temp. 20.5C
Taubaté, Brasil
Mean temp. 20.4C
Teixeira de Freitas, Brasil
Mean temp. 23.9C
Teresina, Brasil
Mean temp. 27.8C
Teresópolis, Brasil
Mean temp. 18.7C
Teófilo Otoni, Brasil
Mean temp. 22.6C
Toledo, Brasil
Mean temp. 21C
Tubarao, Brasil
Mean temp. 20C
Uberaba, Brasil
Mean temp. 22.7C
Uberlândia, Brasil
Mean temp. 22.2C
Uruguaiana, Brasil
Mean temp. 19.9C
Varginha, Brasil
Mean temp. 19.9C
RR
2.00
1.50
1.25
1.00
0.80
Temperature (°C)

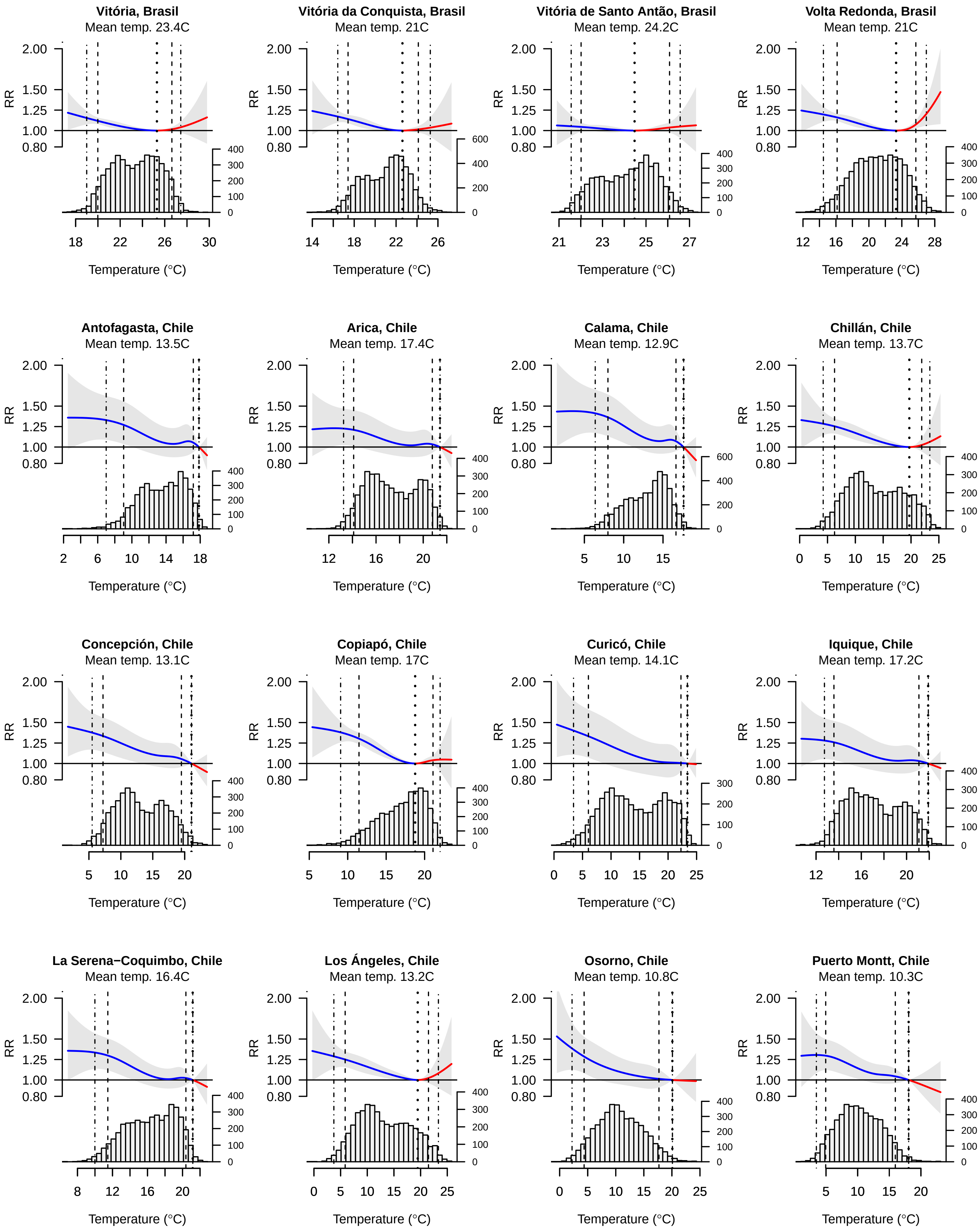

Vitória, Brasil
Mean temp. 23.4C
Vitória da Conquista, Brasil
Mean temp. 21C
Vitória de Santo Antão, Brasil
Mean temp. 24.2C
Volta Redonda, Brasil
Mean temp. 21C
Antofagasta, Chile
Mean temp. 13.5C
Arica, Chile
Mean temp. 17.4C
Calama, Chile
Mean temp. 12.9C
Chillán, Chile
Mean temp. 13.7C
Concepción, Chile
Mean temp. 13.1C
Copiapó, Chile
Mean temp. 17C
Curicó, Chile
Mean temp. 14.1C
Iquique, Chile
Mean temp. 17.2C
La Serena−Coquimbo, Chile
Mean temp. 16.4C
Los Ángeles, Chile
Mean temp. 13.2C
Osorno, Chile
Mean temp. 10.8C
Puerto Montt, Chile
Mean temp. 10.3C
RR
Temperature (°C)

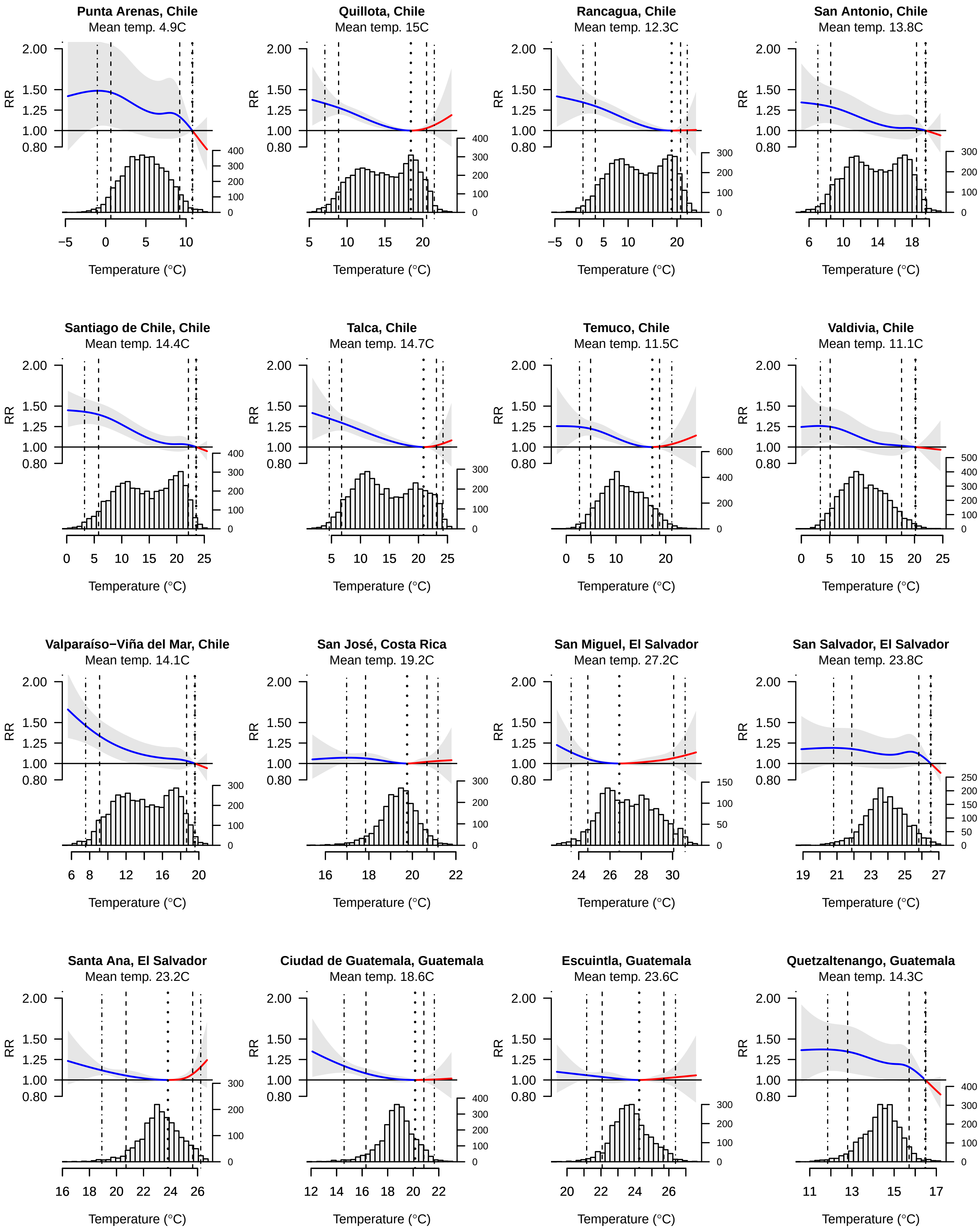
Punta Arenas, Chile
Mean temp. 4.9C
Quillota, Chile
Mean temp. 15C
Rancagua, Chile
Mean temp. 12.3C
San Antonio, Chile
Mean temp. 13.8C
Santiago de Chile, Chile
Mean temp. 14.4C
Talca, Chile
Mean temp. 14.7C
Temuco, Chile
Mean temp. 11.5C
Valdivia, Chile
Mean temp. 11.1C
Valparaíso−Viña del Mar, Chile
Mean temp. 14.1C
San José, Costa Rica
Mean temp. 19.2C
San Miguel, El Salvador
Mean temp. 27.2C
San Salvador, El Salvador
Mean temp. 23.8C
Santa Ana, El Salvador
Mean temp. 23.2C
Ciudad de Guatemala, Guatemala
Mean temp. 18.6C
Escuintla, Guatemala
Mean temp. 23.6C
Quetzaltenango, Guatemala
Mean temp. 14.3C
RR
Temperature (°C)

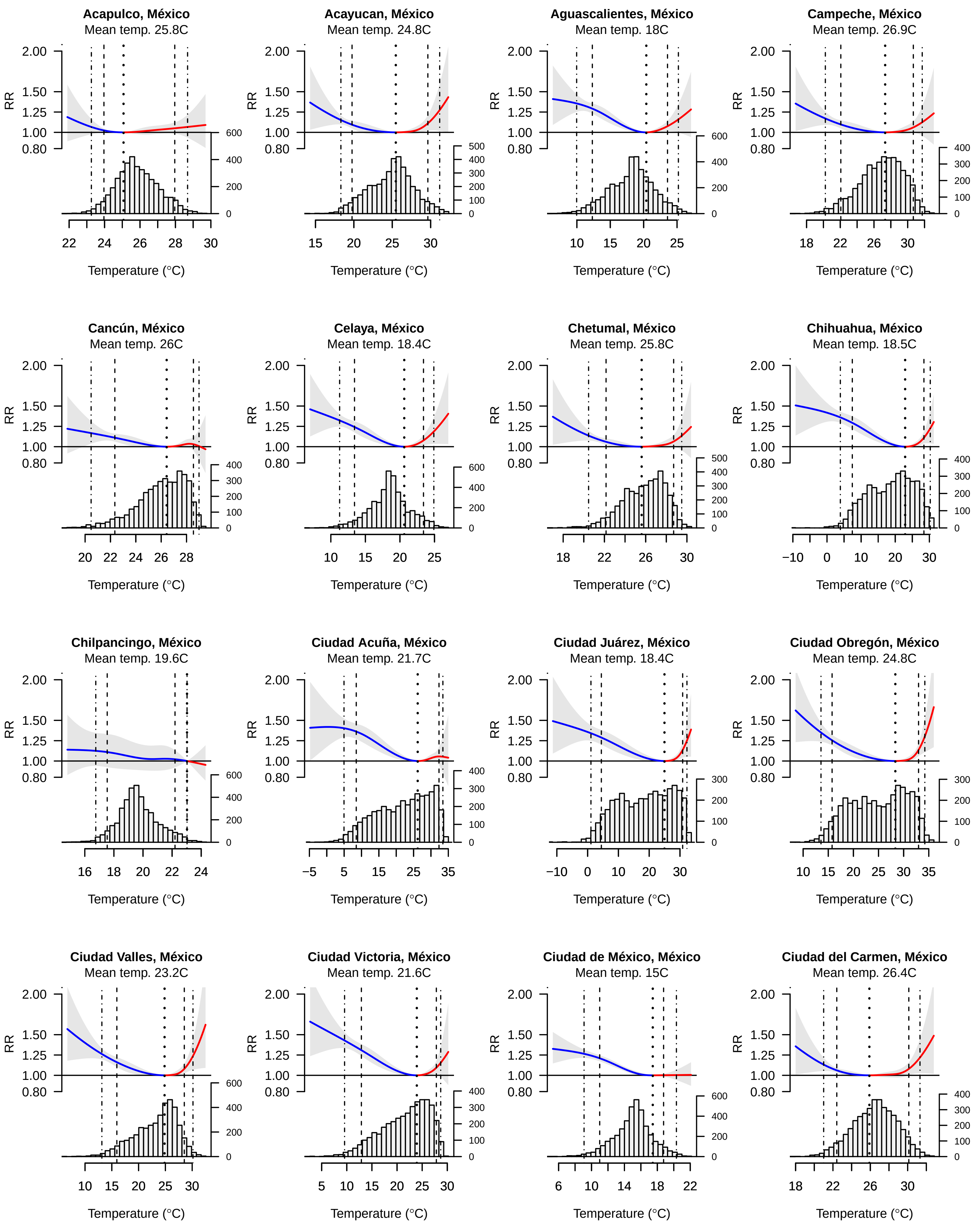
Acapulco, México
Mean temp. 25.8C
Acayucan, México
Mean temp. 24.8C
Aguascalientes, México
Mean temp. 18C
Campeche, México
Mean temp. 26.9C
Cancún, México
Mean temp. 26C
Celaya, México
Mean temp. 18.4C
Chetumal, México
Mean temp. 25.8C
Chihuahua, México
Mean temp. 18.5C
Chilpancingo, México
Mean temp. 19.6C
Ciudad Acuña, México
Mean temp. 21.7C
Ciudad Juárez, México
Mean temp. 18.4C
Ciudad Obregón, México
Mean temp. 24.8C
Ciudad Valles, México
Mean temp. 23.2C
Ciudad Victoria, México
Mean temp. 21.6C
Ciudad de México, México
Mean temp. 15C
Ciudad del Carmen, México
Mean temp. 26.4C
RR
Temperature (°C)

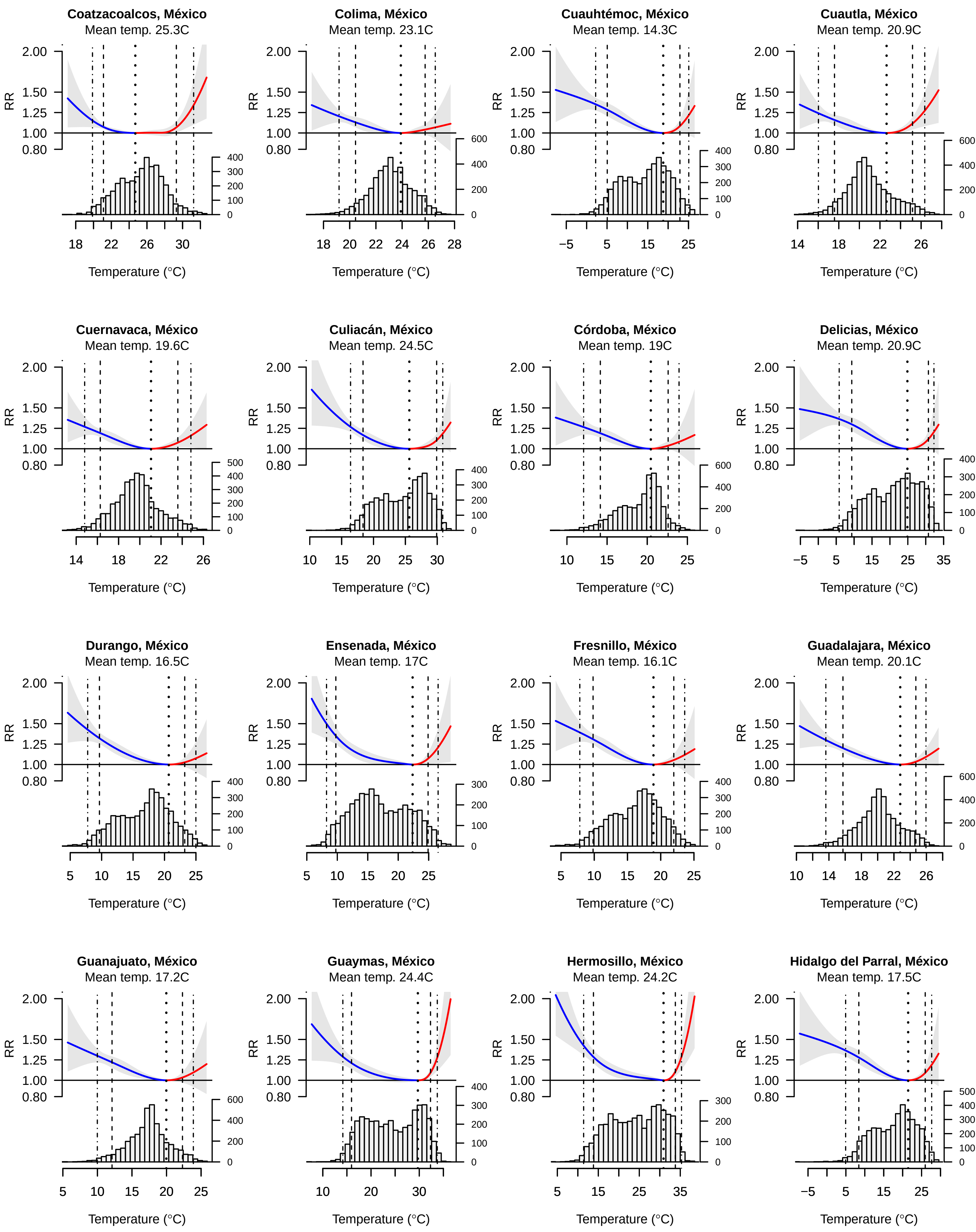
Coatzacoalcos, México
Mean temp. 25.3C
Colima, México
Mean temp. 23.1C
Cuauhtémoc, México
Mean temp. 14.3C
Cuautla, México
Mean temp. 20.9C
Cuernavaca, México
Mean temp. 19.6C
Culiacán, México
Mean temp. 24.5C
Córdoba, México
Mean temp. 19C
Delicias, México
Mean temp. 20.9C
Durango, México
Mean temp. 16.5C
Ensenada, México
Mean temp. 17C
Fresnillo, México
Mean temp. 16.1C
Guadalajara, México
Mean temp. 20.1C
Guanajuato, México
Mean temp. 17.2C
Guaymas, México
Mean temp. 24.4C
Hermosillo, México
Mean temp. 24.2C
Hidalgo del Parral, México
Mean temp. 17.5C
RR
Temperature (°C)

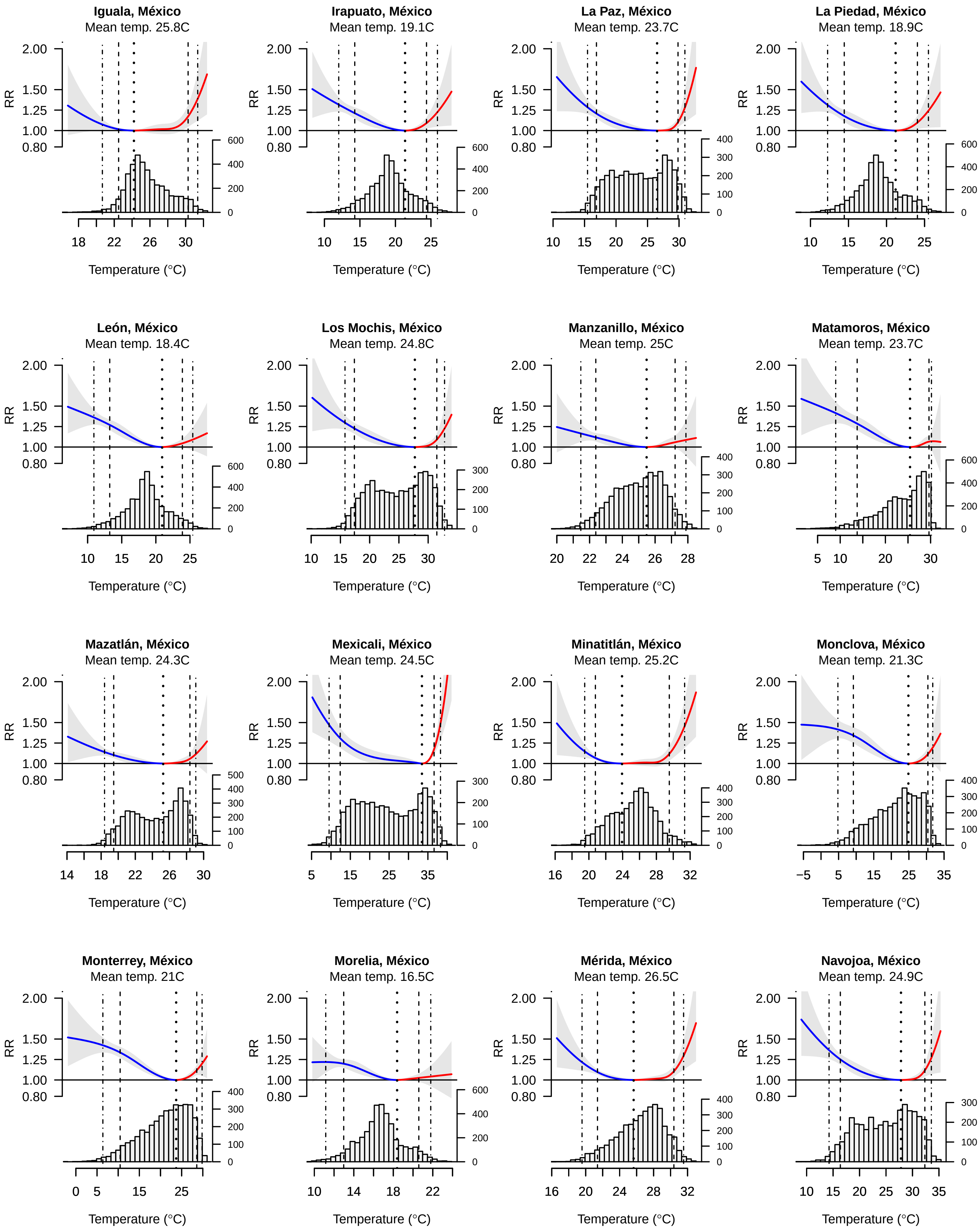
Iguala, México
Mean temp. 25.8C
Irapuato, México
Mean temp. 19.1C
La Paz, México
Mean temp. 23.7C
La Piedad, México
Mean temp. 18.9C
León, México
Mean temp. 18.4C
Los Mochis, México
Mean temp. 24.8C
Manzanillo, México
Mean temp. 25C
Matamoros, México
Mean temp. 23.7C
Mazatlán, México
Mean temp. 24.3C
Mexicali, México
Mean temp. 24.5C
Minatitlán, México
Mean temp. 25.2C
Monclova, México
Mean temp. 21.3C
Monterrey, México
Mean temp. 21C
Morelia, México
Mean temp. 16.5C
Mérida, México
Mean temp. 26.5C
Navojoa, México
Mean temp. 24.9C
RR
Temperature (°C)

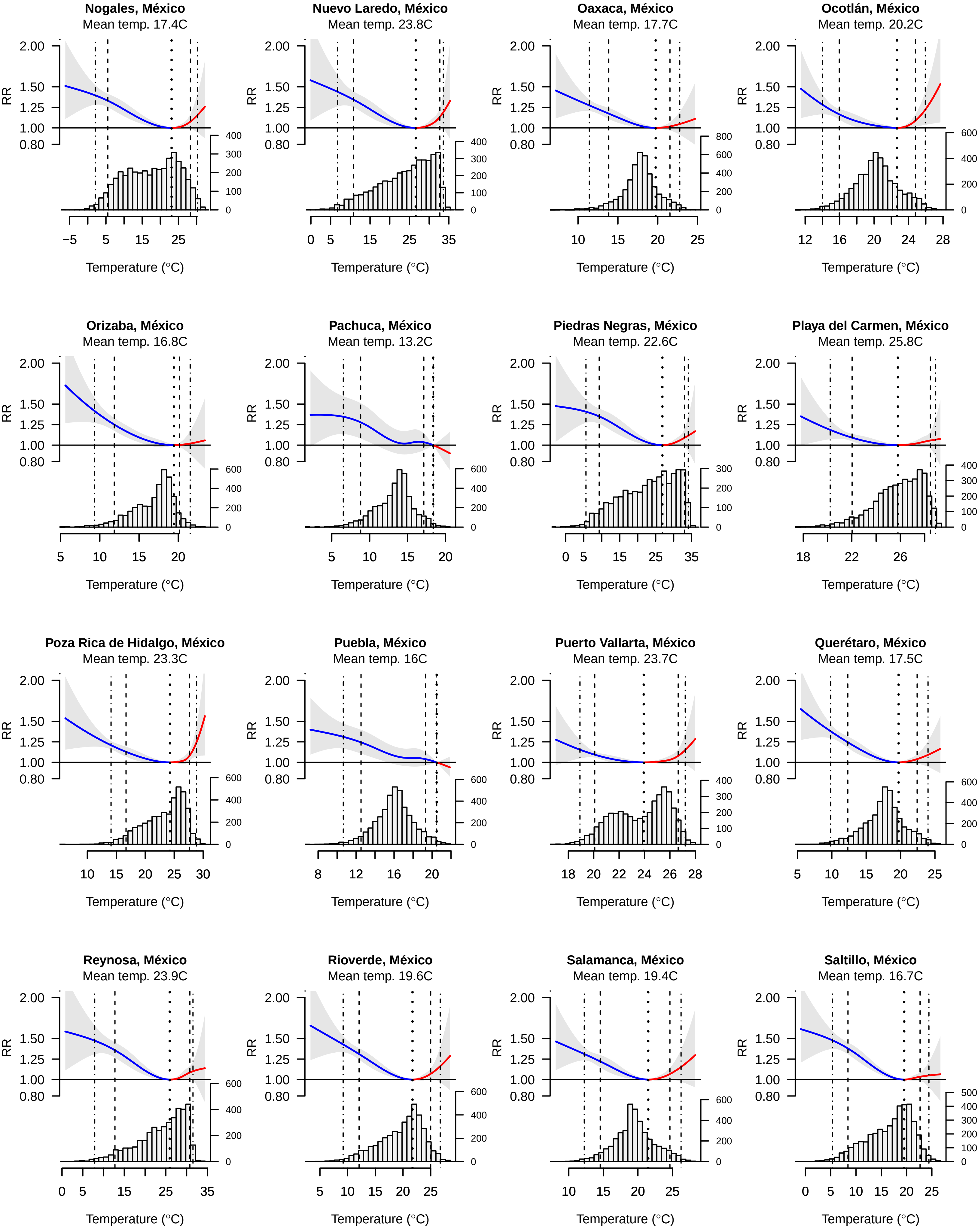

Nogales, México
Mean temp. 17.4C
Nuevo Laredo, México
Mean temp. 23.8C
Oaxaca, México
Mean temp. 17.7C
Ocotlán, México
Mean temp. 20.2C
Orizaba, México
Mean temp. 16.8C
Pachuca, México
Mean temp. 13.2C
Piedras Negras, México
Mean temp. 22.6C
Playa del Carmen, México
Mean temp. 25.8C
Poza Rica de Hidalgo, México
Mean temp. 23.3C
Puebla, México
Mean temp. 16C
Puerto Vallarta, México
Mean temp. 23.7C
Querétaro, México
Mean temp. 17.5C
Reynosa, México
Mean temp. 23.9C
Rioverde, México
Mean temp. 19.6C
Salamanca, México
Mean temp. 19.4C
Saltillo, México
Mean temp. 16.7C
RR
2.00
1.50
1.25
1.00
0.80
Temperature (°C)

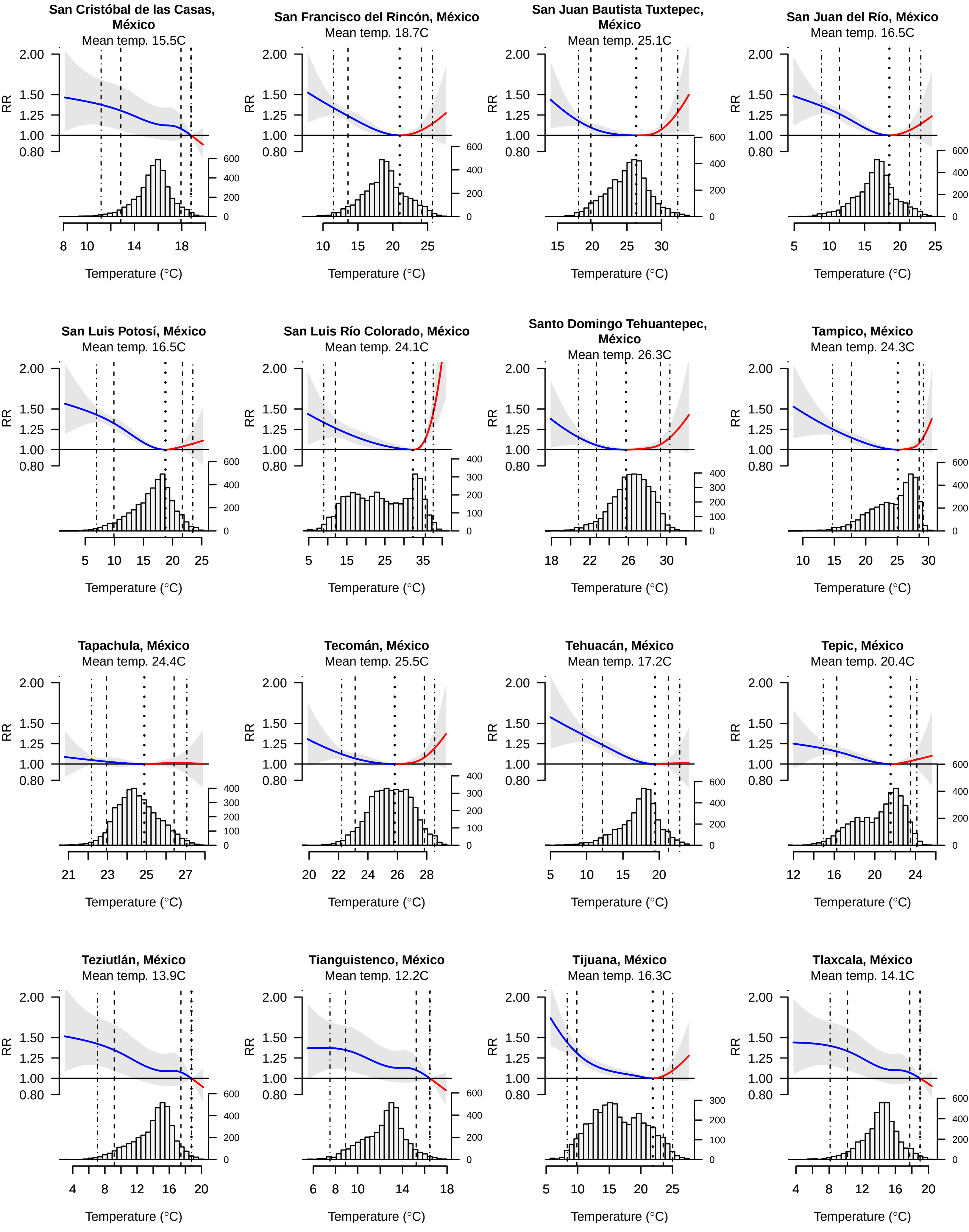

San Cristóbal de las Casas, México
Mean temp. 15.5C
San Francisco del Rincón, México
Mean temp. 18.7C
San Juan Bautista Tuxtepec, México
Mean temp. 25.1C
San Juan del Río, México
Mean temp. 16.5C
San Luis Potosí, México
Mean temp. 16.5C
San Luis Río Colorado, México
Mean temp. 24.1C
Santo Domingo Tehuantepec, México
Mean temp. 26.3C
Tampico, México
Mean temp. 24.3C
Tapachula, México
Mean temp. 24.4C
Tecomán, México
Mean temp. 25.5C
Tehuacán, México
Mean temp. 17.2C
Tepic, México
Mean temp. 20.4C
Teziutlán, México
Mean temp. 13.9C
Tianguistenco, México
Mean temp. 12.2C
Tijuana, México
Mean temp. 16.3C
Tlaxcala, México
Mean temp. 14.1C
RR
2.00
1.50
1.25
1.00
0.80
Temperature (°C)

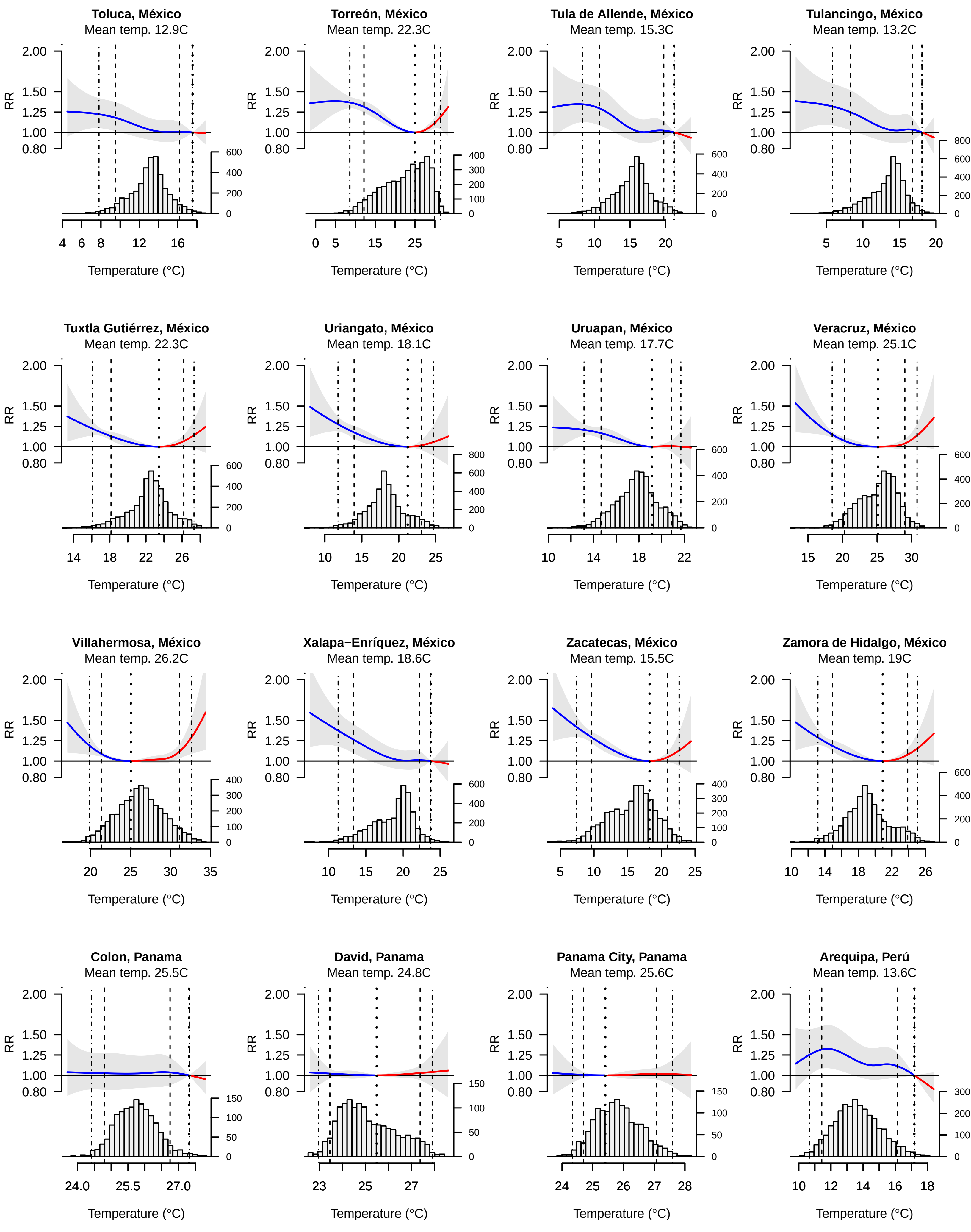
Toluca, México
Mean temp. 12.9C
Torreón, México
Mean temp. 22.3C
Tula de Allende, México
Mean temp. 15.3C
Tulancingo, México
Mean temp. 13.2C
Tuxtla Gutiérrez, México
Mean temp. 22.3C
Uriangato, México
Mean temp. 18.1C
Uruapan, México
Mean temp. 17.7C
Veracruz, México
Mean temp. 25.1C
Villahermosa, México
Mean temp. 26.2C
Xalapa−Enríquez, México
Mean temp. 18.6C
Zacatecas, México
Mean temp. 15.5C
Zamora de Hidalgo, México
Mean temp. 19C
Colon, Panama
Mean temp. 25.5C
David, Panama
Mean temp. 24.8C
Panama City, Panama
Mean temp. 25.6C
Arequipa, Perú
Mean temp. 13.6C
RR
2.00
1.50
1.25
1.00
0.80
Temperature (°C)

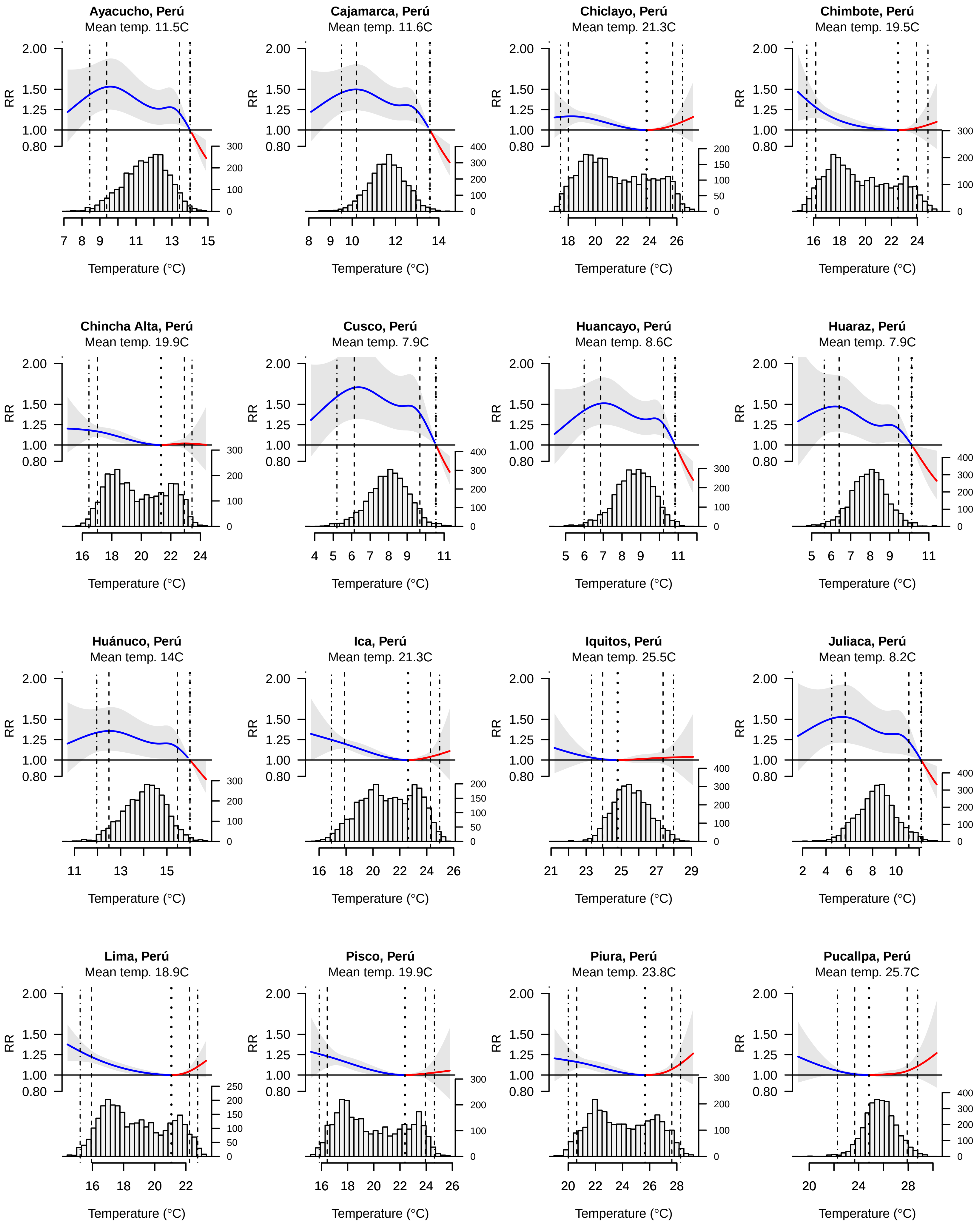
Ayacucho, Perú
Mean temp. 11.5C
Cajamarca, Perú
Mean temp. 11.6C
Chiclayo, Perú
Mean temp. 21.3C
Chimbote, Perú
Mean temp. 19.5C
Chincha Alta, Perú
Mean temp. 19.9C
Cusco, Perú
Mean temp. 7.9C
Huancayo, Perú
Mean temp. 8.6C
Huaraz, Perú
Mean temp. 7.9C
Huánuco, Perú
Mean temp. 14C
Ica, Perú
Mean temp. 21.3C
Iquitos, Perú
Mean temp. 25.5C
Juliaca, Perú
Mean temp. 8.2C
Lima, Perú
Mean temp. 18.9C
Pisco, Perú
Mean temp. 19.9C
Piura, Perú
Mean temp. 23.8C
Pucallpa, Perú
Mean temp. 25.7C
RR
Temperature (°C)

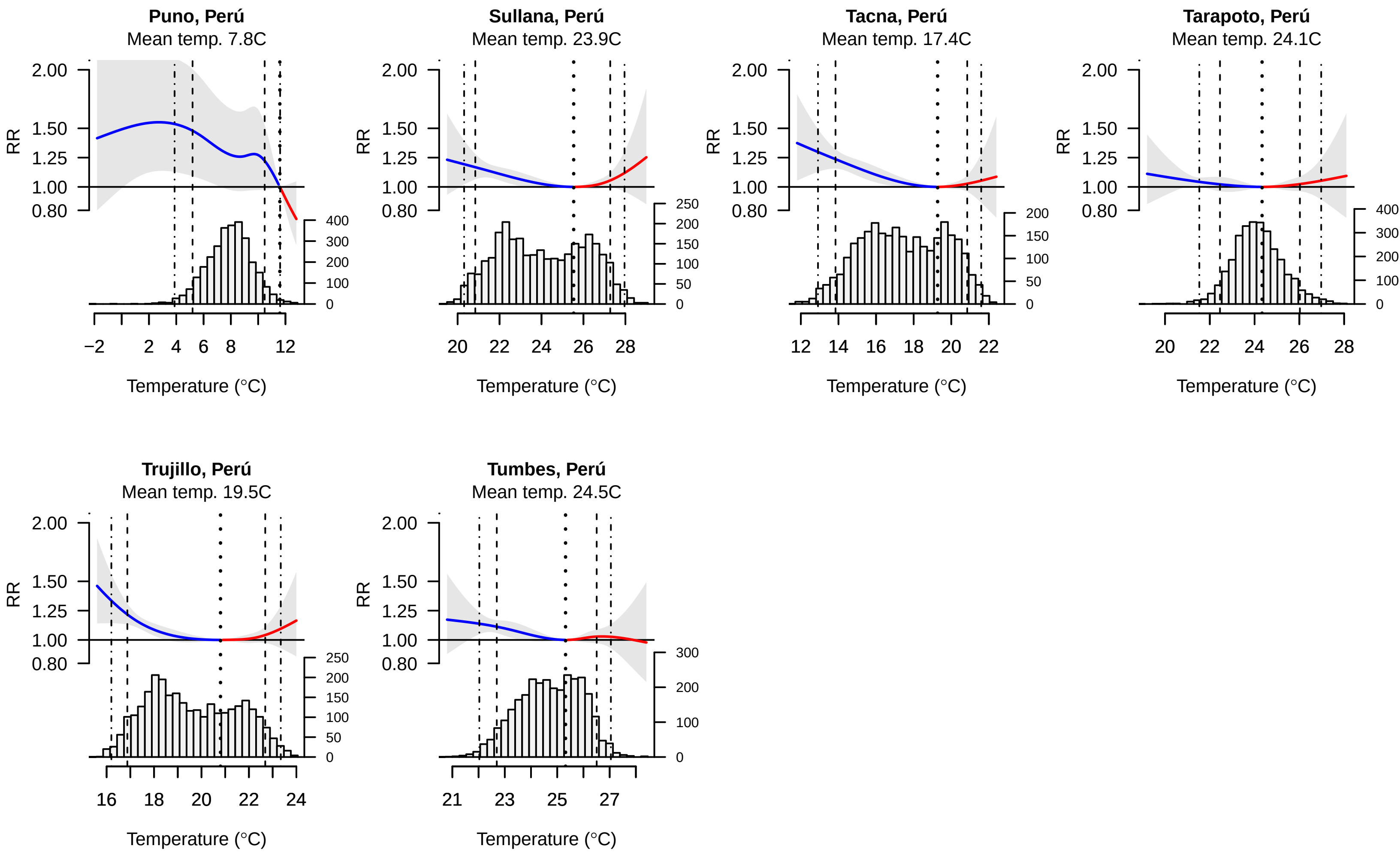

Puno, Perú
Mean temp. 7.8C
Sullana, Perú
Mean temp. 23.9C
Tacna, Perú
Mean temp. 17.4C
Tarapoto, Perú
Mean temp. 24.1C
Trujillo, Perú
Mean temp. 19.5C
Tumbes, Perú
Mean temp. 24.5C
RR
2.00
1.50
1.25
1.00
0.80
Temperature (°C)

**Fig. S1**. City-specific temperature-mortality relationship for all-cause, all-ages mortality.

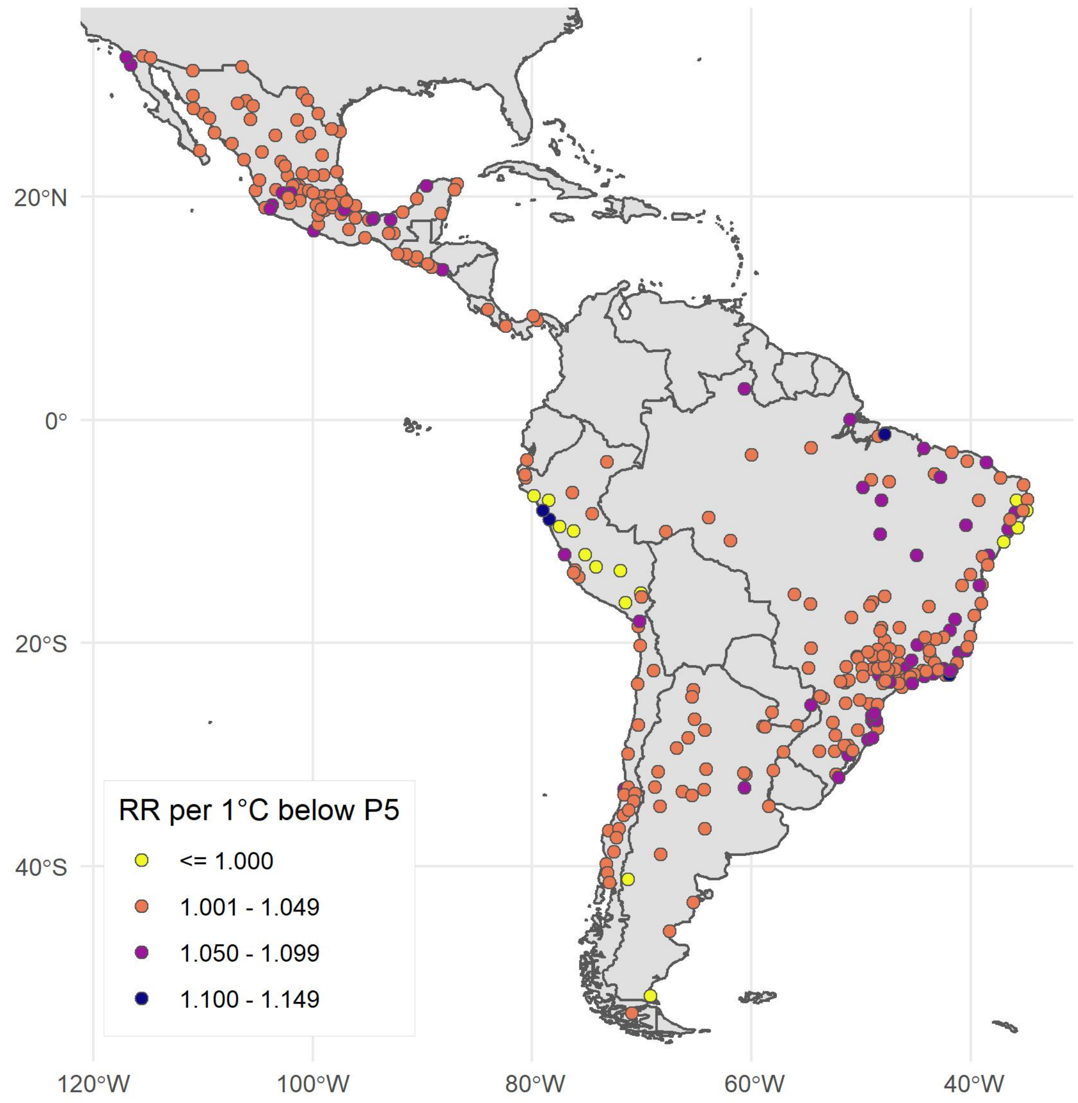


**Fig. S2.** City-specific change in relative risk of mortality per 1°C decrease in temperature below 5th percentile observed daily temperature

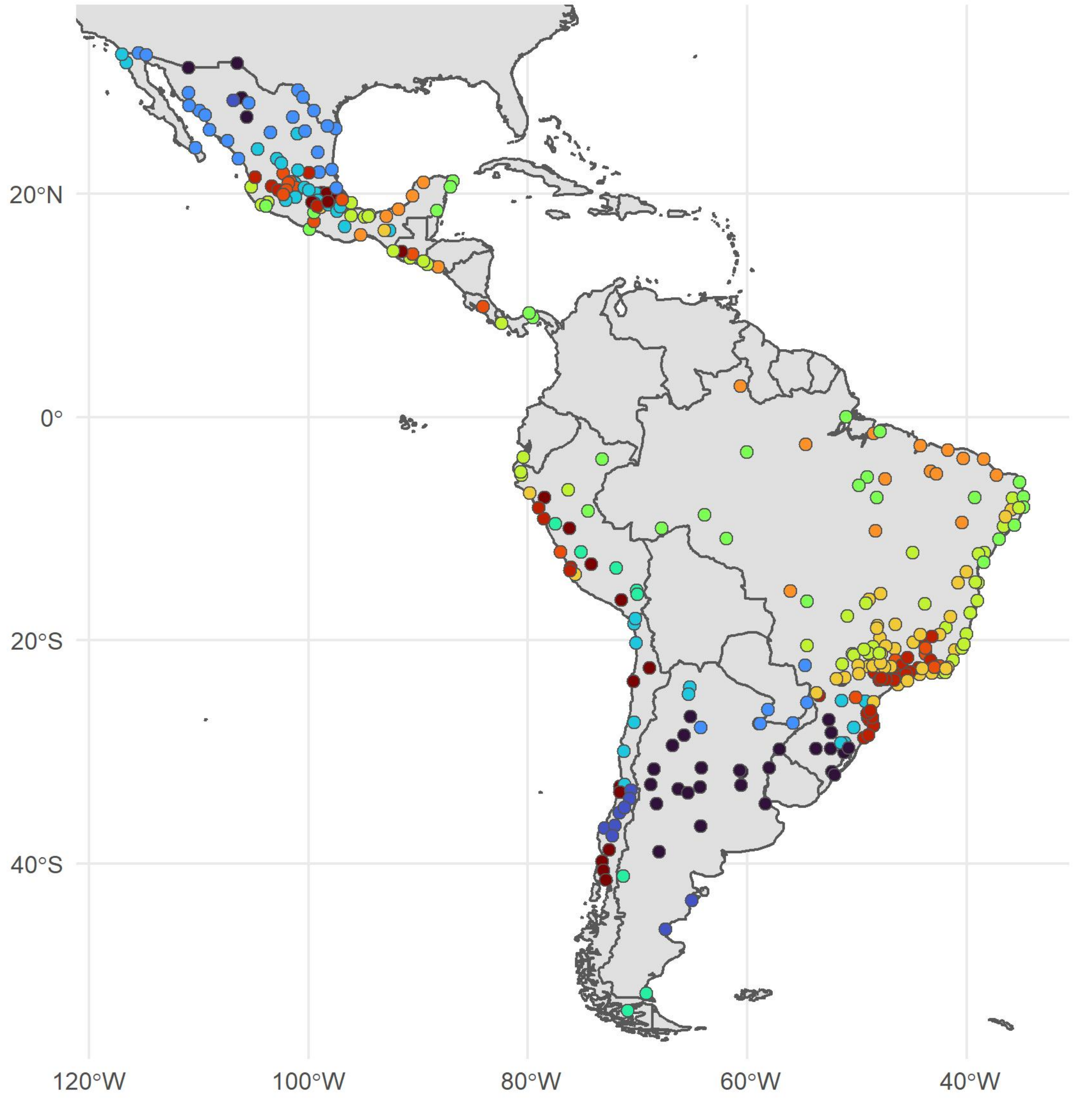


**Fig. S3.** Cluster groupings in analysis of cause-specific mortality outcomes (by temperature, N=12 clusters).

**Table S2.** City-specific daily temperature distributions and temperature-mortality associations

| | | | Temperature | | | Temperature Association with Mortality | | |
|---|---|---|---|---|---|---|---|---|
| Country | City | Total Number of Deaths | 5th Percentile Temperature | Median Temperature | 95th Percentile Temperature | Minimum Mortality Temperature ⁰C | Excess Death Fraction All Cold (%) | Excess Death Fraction All Heat (%) |
| **Argentina** | Buenos Aires | 172311 | 7.9 | 17.0 | 25.9 | 23.5 | 15.2 | 2 |
| | Comodoro Rivadavia | 7682 | 3.6 | 11.8 | 21.0 | 18 | 5.8 | -1 |
| | Concordia | 8562 | 8.9 | 19.2 | 27.5 | 24.8 | 8.2 | 1.2 |
| | Corrientes | 16278 | 11.7 | 22.1 | 29.4 | 26.2 | 17.7 | 1.2 |
| | Córdoba | 84196 | 7.9 | 17.7 | 25.7 | 22.2 | 6.6 | 1.5 |
| | Formosa | 10354 | 12.4 | 23.4 | 30.5 | 26.7 | 16.4 | -1.6 |
| | La Rioja | 7363 | 8.7 | 19.4 | 28.7 | 24.6 | 11.2 | 2.4 |
| | Mendoza | 57175 | 5.3 | 15.9 | 26.0 | 22.3 | 11.5 | 1.6 |
| | Neuquén-Plottier-Cipolletti | 28411 | 5.3 | 15.3 | 27.0 | 23.2 | 9.5 | 0.7 |
| | Paraná | 18864 | 8.8 | 19.0 | 27.3 | 24.7 | 8.1 | 1.5 |
| | Posadas | 14997 | 12.0 | 21.9 | 28.2 | 25.4 | 9.6 | 2 |
| | Rawson - Trelew | 5745 | 5.2 | 14.2 | 24.4 | 21 | 8.5 | 0.6 |
| | Resistencia | 18941 | 11.6 | 22.2 | 29.8 | 26.6 | 14.6 | -0.3 |
| | Rosario | 90402 | 8.5 | 18.5 | 26.9 | 24.5 | 10.7 | 1.4 |
| | Río Cuarto | 16680 | 7.9 | 17.6 | 26.5 | 22.9 | 3 | 2.1 |
| | Río Gallegos | 3092 | -0.4 | 6.8 | 14.4 | 16.7 | -23.8 | 0.1 |
| | Salta | 24357 | 7.9 | 16.5 | 22.1 | 19 | 5.2 | 0.7 |
| | San Carlos de Bariloche | 5002 | -1.2 | 5.2 | 16.8 | 18.8 | -4.8 | 0.3 |

| | | | Temperature | | | Temperature Association with Mortality | | |
|---|---|---|---|---|---|---|---|---|
| Country | City | Total Number of Deaths | 5th Percentile Temperature | Median Temperature | 95th Percentile Temperature | Minimum Mortality Temperature $^0$C | Excess Death Fraction All Cold (%) | Excess Death Fraction All Heat (%) |
| | San Fernando del Valle de Catamarca | 8270 | 8.6 | 19.6 | 28.1 | 24.3 | 7.5 | -0.8 |
| | San Juan | 24962 | 7.3 | 17.9 | 26.9 | 22.6 | 6.3 | 1.5 |
| | San Luis | 8448 | 7.3 | 17.8 | 27.7 | 24.4 | 29.7 | 1.4 |
| | San Miguel de Tucumán-Tafí Viejo | 47772 | 10.0 | 19.1 | 25.9 | 23.8 | 8.9 | 1.8 |
| | San Rafael | 10313 | 5.5 | 16.4 | 27.3 | 23.2 | -2.9 | 1.5 |
| | San Salvador de Jujuy | 13011 | 8.0 | 15.6 | 20.6 | 17.7 | 2.5 | 0.6 |
| | Santa Fe | 31557 | 9.4 | 19.5 | 27.8 | 25.1 | 6.7 | 1.5 |
| | Santa Rosa-Toay | 5910 | 6.5 | 16.5 | 27.0 | 23.8 | 15.5 | 2 |
| | Santiago del Estero- La Banda | 19492 | 12.4 | 22.7 | 31.1 | 27.3 | 3.2 | 1.2 |
| | Villa Mercedes | 6426 | 7.4 | 17.4 | 26.9 | 23.9 | -9.7 | 0.6 |
| **Brasil** | Alagoinhas | 12167 | 21.5 | 24.7 | 27.5 | 25.2 | 1.8 | -1.2 |
| | Angra dos Reis | 11764 | 16.8 | 21.6 | 26.2 | 24.6 | -1.1 | -0.7 |
| | Anápolis | 28352 | 19.7 | 22.0 | 25.8 | 23.8 | 2.2 | -0.2 |
| | Apucarana | 10514 | 14.1 | 21.0 | 24.8 | 22.5 | 3 | 2.3 |
| | Aracaju | 58215 | 23.2 | 25.5 | 27.3 | 25.9 | 3.6 | 0.6 |
| | Araguari | 10117 | 18.8 | 22.4 | 26.0 | 23.9 | -10.5 | 0.9 |
| | Araguaína | 9928 | 24.2 | 26.3 | 29.1 | 25.8 | 3 | 6 |
| | Arapiraca | 17849 | 21.6 | 24.9 | 27.5 | 25.7 | 5.1 | -2.2 |
| | Arapongas | 14252 | 14.3 | 21.2 | 25.0 | 23.1 | 3.4 | 0.5 |
| | Araraquara | 22801 | 17.3 | 22.5 | 26.2 | 23.8 | -0.3 | 0.6 |

| | | | Temperature | | | Temperature Association with Mortality | | |
|---|---|---|---|---|---|---|---|---|
| Country | City | Total Number of Deaths | 5th Percentile Temperature | Median Temperature | 95th Percentile Temperature | Minimum Mortality Temperature $^0$C | Excess Death Fraction All Cold (%) | Excess Death Fraction All Heat (%) |
| | Araras | 10922 | 16.1 | 21.7 | 25.6 | 23.7 | 0.4 | 2 |
| | Araruama | 21423 | 19.4 | 23.0 | 26.7 | 24.9 | 11.1 | 1.7 |
| | Araçatuba | 18892 | 18.7 | 24.4 | 28.2 | 25 | 1.2 | 3.7 |
| | Atibaia | 13875 | 14.0 | 19.6 | 23.7 | 21.9 | 5.6 | 2.4 |
| | Balneário Camboriú | 10988 | 14.4 | 20.5 | 25.8 | 23.9 | -2.3 | -1.1 |
| | Barbacena | 12760 | 13.7 | 18.7 | 22.2 | 20.9 | 11.4 | 0.2 |
| | Barreiras | 7798 | 22.3 | 24.8 | 28.9 | 24.2 | 1.2 | 4.8 |
| | Barretos | 13154 | 19.1 | 23.8 | 27.6 | 25.2 | 7.1 | 1.8 |
| | Bauru | 32505 | 16.8 | 22.6 | 26.6 | 24 | 3.6 | 1.9 |
| | Belo Horizonte | 357006 | 16.4 | 20.9 | 24.3 | 22 | 2 | 0.6 |
| | Belém | 150465 | 24.9 | 26.6 | 28.3 | 28.7 | 8.4 | 0 |
| | Bento Goncalves | 12198 | 8.7 | 17.8 | 23.5 | 21.3 | 16.7 | 0.9 |
| | Birigui | 9026 | 18.6 | 24.3 | 28.1 | 25.4 | 6.3 | 1.9 |
| | Blumenau | 34585 | 13.1 | 19.7 | 25.2 | 23.1 | 0.5 | 0.4 |
| | Boa Vista | 14681 | 24.4 | 26.7 | 29.8 | 26.3 | -0.3 | 2.1 |
| | Botucatu | 11829 | 14.7 | 20.7 | 24.6 | 23.3 | 2.9 | 0.4 |
| | Bragança Paulista | 14621 | 14.2 | 19.9 | 23.9 | 22.3 | 9.7 | 1.4 |
| | Brasília | 178255 | 19.0 | 21.6 | 25.4 | 23.4 | 2 | 0.6 |
| | Brusque | 8200 | 12.9 | 19.6 | 25.1 | 23.3 | -2.5 | -1.4 |
| | Cabo Frio | 24696 | 19.6 | 23.0 | 26.4 | 25.5 | 9.1 | 0.4 |

| | | | Temperature | | | Temperature Association with Mortality | | |
|---|---|---|---|---|---|---|---|---|
| Country | City | Total Number of Deaths | 5th Percentile Temperature | Median Temperature | 95th Percentile Temperature | Minimum Mortality Temperature $^{0}C$ | Excess Death Fraction All Cold (%) | Excess Death Fraction All Heat (%) |
| | Cachoeiro de Itapemirim | 16980 | 18.1 | 22.5 | 27.1 | 25.3 | 4.8 | 1.3 |
| | Campina Grande | 40634 | 21.1 | 23.7 | 25.4 | 25.9 | 7.4 | 0 |
| | Campinas | 219551 | 15.8 | 21.4 | 25.5 | 24 | 6.3 | 0.6 |
| | Campo Grande | 59982 | 17.6 | 24.1 | 27.6 | 25.7 | 4 | -0.5 |
| | Campos dos Goytacazes | 49472 | 19.7 | 23.8 | 27.8 | 25.6 | 4.4 | 0.7 |
| | Caraguatatuba | 13929 | 16.8 | 21.4 | 26.1 | 24.1 | -5.6 | 2.6 |
| | Caruaru | 27763 | 20.2 | 23.0 | 25.0 | 23.8 | 5 | 0.5 |
| | Cascavel | 20823 | 12.3 | 20.9 | 24.9 | 22.9 | 4.8 | 0.4 |
| | Castanhal | 11036 | 24.5 | 26.2 | 28.5 | 25.8 | 6.2 | 6.6 |
| | Catanduva | 13262 | 18.3 | 23.5 | 27.2 | 24.5 | 2 | 3.2 |
| | Caxias | 10829 | 24.5 | 27.5 | 30.9 | 26.9 | -0.1 | 0 |
| | Caxias do Sul | 32310 | 8.4 | 17.4 | 22.8 | 19.7 | 5.2 | -0.3 |
| | Chapecó | 11170 | 10.6 | 19.8 | 24.4 | 22.2 | 7.4 | -0.1 |
| | Conselheiro Lafaiete | 9597 | 14.6 | 19.5 | 22.9 | 21.5 | -4.6 | -0.6 |
| | Criciúma | 23285 | 12.7 | 19.9 | 25.6 | 23.7 | 2.3 | 1.2 |
| | Cuiabá | 60535 | 22.0 | 26.5 | 30.0 | 26.4 | -0.5 | 0.6 |
| | Curitiba | 223491 | 10.8 | 17.5 | 22.5 | 19.9 | 6.3 | 0.3 |
| | Divinópolis | 16391 | 17.2 | 21.5 | 25.1 | 23.8 | 10.5 | 0.1 |
| | Dourados | 15753 | 15.9 | 24.3 | 28.3 | 24.8 | 2.9 | 0.1 |
| | Feira de Santana | 42793 | 21.1 | 24.6 | 27.8 | 25.3 | 2.4 | 1.1 |

| | | | Temperature | | | Temperature Association with Mortality | | |
|---|---|---|---|---|---|---|---|---|
| Country | City | Total Number of Deaths | 5th Percentile Temperature | Median Temperature | 95th Percentile Temperature | Minimum Mortality Temperature $^{0}C$ | Excess Death Fraction All Cold (%) | Excess Death Fraction All Heat (%) |
| | Florianópolis | 55776 | 14.2 | 20.4 | 25.8 | 23.8 | 7.5 | 0.4 |
| | Fortaleza | 246596 | 25.2 | 26.7 | 27.7 | 28.1 | 8.1 | 0 |
| | Foz do Iguaçu | 20321 | 13.6 | 23.1 | 28.0 | 25.5 | 2.1 | -1.1 |
| | Franca | 26382 | 17.1 | 21.6 | 25.0 | 23.5 | 4.5 | 0.1 |
| | Garanhuns | 11735 | 18.9 | 21.9 | 24.3 | 23 | 9.1 | -1.3 |
| | Goiânia | 148759 | 20.8 | 23.2 | 27.3 | 24.6 | 2 | 1.2 |
| | Governador Valadares | 23969 | 20.3 | 24.2 | 28.1 | 23.9 | 1 | 1.7 |
| | Guarapari | 8318 | 19.7 | 23.3 | 26.7 | 24.5 | -8.4 | -2.5 |
| | Guarapuava | 14754 | 10.3 | 17.9 | 21.8 | 19.7 | -1 | -0.4 |
| | Guaratinguetá | 25657 | 15.4 | 20.8 | 25.1 | 23.2 | 2.4 | -0.2 |
| | Ilhéus | 16926 | 21.2 | 24.0 | 26.2 | 24.4 | 4 | 2.8 |
| | Imperatriz | 21849 | 24.5 | 26.8 | 29.8 | 27.4 | 1.2 | -2.7 |
| | Ipatinga | 38200 | 18.8 | 22.8 | 26.7 | 24.4 | 6.8 | 1.3 |
| | Itabira | 8137 | 16.1 | 20.5 | 23.9 | 22.3 | 6.2 | -0.5 |
| | Itabuna | 21001 | 20.7 | 23.6 | 25.8 | 25.1 | 10.8 | 0.4 |
| | Itajaí | 23982 | 14.8 | 20.8 | 26.0 | 24.3 | -1.2 | 0.4 |
| | Itapetininga | 13360 | 14.0 | 20.1 | 24.4 | 22.7 | 9.9 | 0.2 |
| | Jaraguá do Sul | 11859 | 13.6 | 20.1 | 25.6 | 23.6 | 13.1 | 0.4 |
| | Jaú | 13276 | 16.7 | 22.4 | 26.5 | 23.7 | 2.1 | 3.8 |
| | Jequié | 13364 | 19.4 | 23.0 | 25.7 | 23.5 | -3.1 | 1 |

| | | | Temperature | | | Temperature Association with Mortality | | |
|---|---|---|---|---|---|---|---|---|
| Country | City | Total Number of Deaths | 5th Percentile Temperature | Median Temperature | 95th Percentile Temperature | Minimum Mortality Temperature $^{0}C$ | Excess Death Fraction All Cold (%) | Excess Death Fraction All Heat (%) |
| | Ji-Paraná | 8421 | 23.8 | 25.7 | 28.8 | 26.2 | 5.2 | -0.6 |
| | Joinville | 35248 | 14.4 | 20.8 | 26.2 | 24.2 | 1.6 | 1.2 |
| | João Pessoa | 83786 | 23.9 | 25.8 | 27.3 | 25.5 | 0.2 | 1.3 |
| | Juazeiro do Norte | 34505 | 23.1 | 25.6 | 29.2 | 26.8 | 1.6 | 0.4 |
| | Juiz de Fora | 52045 | 15.3 | 20.3 | 24.3 | 22.9 | 4.3 | -0.1 |
| | Jundiaí | 48893 | 14.2 | 19.9 | 23.9 | 21.9 | 4.4 | 1.2 |
| | Lages (Lajes) | 14922 | 8.3 | 16.4 | 21.6 | 19.1 | 0.8 | 0.6 |
| | Limeira | 25482 | 16.2 | 22.0 | 26.0 | 24.4 | 9 | 0.7 |
| | Linhares | 10155 | 20.6 | 24.0 | 27.0 | 25.1 | 6.9 | 1.8 |
| | Londrina | 55611 | 14.9 | 21.7 | 25.6 | 23 | 4.3 | 1.6 |
| | Macapá | 25510 | 24.4 | 26.3 | 28.5 | 25.8 | 0.9 | 2.4 |
| | Macaé | 14119 | 19.0 | 23.0 | 27.1 | 24.1 | -3.1 | 0.4 |
| | Maceió | 87910 | 23.2 | 25.3 | 27.0 | 25.7 | -0.2 | 0 |
| | Manaus | 114662 | 24.8 | 26.2 | 28.8 | 25.4 | -0.1 | -0.6 |
| | Marabá | 15787 | 24.2 | 26.3 | 29.0 | 27.1 | 12.2 | -2 |
| | Maringá | 38724 | 15.1 | 22.6 | 26.6 | 23.9 | 1.9 | 0.8 |
| | Marília | 19942 | 17.0 | 22.9 | 26.8 | 24.2 | 5.4 | 1.5 |
| | Mogi Guaçu (Moji Guaçu) | 21683 | 16.3 | 21.8 | 25.7 | 23.6 | 7.2 | 0.9 |
| | Montes Claros | 25378 | 19.4 | 23.0 | 27.1 | 25.1 | 1.9 | 1.7 |
| | Mossoró | 18513 | 25.7 | 27.9 | 29.3 | 27.3 | -0.5 | 1.8 |

| | | | Temperature | | | Temperature Association with Mortality | | |
|---|---|---|---|---|---|---|---|---|
| Country | City | Total Number of Deaths | 5th Percentile Temperature | Median Temperature | 95th Percentile Temperature | Minimum Mortality Temperature $^0$C | Excess Death Fraction All Cold (%) | Excess Death Fraction All Heat (%) |
| | Natal | 85164 | 24.2 | 26.0 | 27.3 | 25 | 0.8 | 3.5 |
| | Nova Friburgo | 20192 | 13.6 | 18.5 | 22.6 | 21.6 | 19 | 0.8 |
| | Ourinhos | 10196 | 16.4 | 22.7 | 26.9 | 24 | -0.3 | 2.1 |
| | Palmas | 9187 | 24.3 | 26.6 | 30.2 | 26 | 0.2 | 3 |
| | Paranaguá | 12513 | 15.7 | 21.2 | 26.4 | 24.3 | 4.3 | 1.1 |
| | Parauapebas | 7419 | 23.6 | 25.7 | 28.5 | 25.7 | 2.6 | 1.7 |
| | Parnaíba | 11441 | 25.7 | 27.8 | 28.9 | 26.9 | 0.5 | 2.5 |
| | Parobe | 13586 | 10.2 | 19.1 | 25.0 | 22.7 | 13.3 | 0.6 |
| | Passo Fundo | 17382 | 9.2 | 18.7 | 23.8 | 20.7 | 3.1 | -0.4 |
| | Passos | 10035 | 16.8 | 21.7 | 25.2 | 22.9 | 4.3 | 2.4 |
| | Patos de Minas | 10992 | 18.2 | 21.7 | 25.3 | 23.2 | 7.7 | 1.8 |
| | Pelotas | 41495 | 10.1 | 18.8 | 25.1 | 23.2 | 5.6 | -1.2 |
| | Petrolina | 33104 | 23.3 | 26.4 | 29.8 | 27.2 | 0.9 | 1.4 |
| | Petrópolis | 35441 | 14.6 | 19.6 | 23.9 | 22.5 | 5.9 | -0.4 |
| | Piracicaba | 33655 | 16.1 | 22.1 | 26.1 | 24 | 1.9 | 1.5 |
| | Ponta Grossa | 28686 | 11.7 | 18.7 | 23.0 | 21.5 | 9.6 | 0 |
| | Porto Alegre | 350444 | 11.1 | 19.8 | 26.0 | 23.8 | 5.5 | 1.3 |
| | Porto Seguro | 7656 | 21.5 | 24.3 | 26.6 | 24.7 | 3.1 | 0.5 |
| | Porto Velho | 30179 | 24.3 | 25.9 | 28.5 | 24.8 | -0.3 | 4.9 |
| | Pouso Alegre | 9869 | 15.0 | 20.0 | 23.7 | 22.4 | 8.7 | 1.5 |

| | | | Temperature | | | Temperature Association with Mortality | | |
|---|---|---|---|---|---|---|---|---|
| Country | City | Total Number of Deaths | 5th Percentile Temperature | Median Temperature | 95th Percentile Temperature | Minimum Mortality Temperature $^0C$ | Excess Death Fraction All Cold (%) | Excess Death Fraction All Heat (%) |
| | Poços de Caldas | 14490 | 13.6 | 18.6 | 21.8 | 20.6 | 11.3 | 1.3 |
| | Presidente Prudente | 21703 | 17.1 | 23.8 | 27.7 | 24.5 | 3 | 2.9 |
| | Recife | 312731 | 23.5 | 25.6 | 27.3 | 26 | -0.3 | 0.2 |
| | Resende | 14138 | 15.5 | 20.8 | 25.4 | 23.3 | 9.6 | 0.1 |
| | Ribeirão Preto | 51462 | 17.9 | 23.0 | 26.7 | 24 | 1.9 | 2 |
| | Rio Branco | 22061 | 23.0 | 25.4 | 28.0 | 25.8 | 2.8 | 0.9 |
| | Rio Claro | 20396 | 16.2 | 21.8 | 25.8 | 24.4 | 12.9 | -0.2 |
| | Rio Grande | 23032 | 10.9 | 18.8 | 24.8 | 23.1 | -2.3 | 0.6 |
| | Rio Verde | 11892 | 20.0 | 23.3 | 26.8 | 25 | -4.2 | 1.4 |
| | Rio das Ostras | 9353 | 18.8 | 23.0 | 27.2 | 25.3 | 4.7 | -0.2 |
| | Rio de Janeiro | 1282138 | 18.4 | 23.0 | 28.1 | 23.3 | 2.1 | 1.2 |
| | Rondonópolis | 14730 | 22.5 | 26.0 | 29.4 | 26.6 | -0.2 | -2.2 |
| | Salvador | 239702 | 23.0 | 25.3 | 27.3 | 28 | 5.5 | 0 |
| | Santa Cruz do Sul | 11848 | 10.1 | 19.4 | 25.8 | 23.3 | 9.7 | 1.1 |
| | Santa Maria | 25688 | 9.6 | 19.6 | 26.3 | 23.7 | 9.6 | -0.7 |
| | Santarém | 16817 | 24.9 | 26.7 | 29.5 | 26.9 | -1.5 | -0.8 |
| | Santos | 168093 | 16.8 | 21.7 | 26.7 | 23.6 | 1.5 | 1.3 |
| | Sertãozinho | 8653 | 18.4 | 23.4 | 27.3 | 25 | 6.5 | 2 |
| | Sete Lagoas | 16699 | 17.5 | 21.7 | 25.4 | 23.8 | 2.7 | 0.7 |
| | Sobral | 13055 | 24.9 | 27.4 | 29.2 | 27 | 2.7 | -1.6 |

| | | | Temperature | | | Temperature Association with Mortality | | |
|---|---|---|---|---|---|---|---|---|
| Country | City | Total Number of Deaths | 5th Percentile Temperature | Median Temperature | 95th Percentile Temperature | Minimum Mortality Temperature $^0$C | Excess Death Fraction All Cold (%) | Excess Death Fraction All Heat (%) |
| | Sorocaba | 62281 | 14.8 | 20.7 | 24.9 | 23.8 | 6.8 | 0.7 |
| | São Carlos | 20224 | 16.1 | 21.3 | 25.0 | 23.4 | 7.2 | 0.3 |
| | São José do Rio Preto | 45225 | 18.6 | 23.7 | 27.3 | 24.7 | 5.5 | 2.5 |
| | São José dos Campos | 69062 | 14.9 | 20.4 | 24.7 | 22.6 | 7.8 | 0.3 |
| | São Luís | 83401 | 25.2 | 27.0 | 28.0 | 28.3 | 7.7 | -0.1 |
| | São Paulo | 1594830 | 13.8 | 19.6 | 24.1 | 22.4 | 5.6 | 0.6 |
| | Tatuí | 10439 | 15.0 | 21.0 | 25.1 | 23.2 | 5.9 | 2.3 |
| | Taubaté | 38986 | 15.2 | 20.7 | 25.0 | 23.1 | 4.4 | 0.7 |
| | Teixeira de Freitas | 10601 | 20.6 | 24.1 | 27.3 | 24.8 | -7.1 | -1.5 |
| | Teresina | 69439 | 24.7 | 27.9 | 31.5 | 27.5 | 1.5 | 1.5 |
| | Teresópolis | 17477 | 13.9 | 18.9 | 23.0 | 22.1 | 21.5 | 0 |
| | Teófilo Otoni | 13949 | 18.7 | 22.7 | 26.7 | 24.7 | -4.1 | 0.5 |
| | Toledo | 8671 | 13.4 | 22.0 | 26.2 | 23.1 | -1.5 | 3.6 |
| | Tubarao | 11146 | 13.3 | 20.3 | 26.3 | 24.3 | 17.7 | 0.4 |
| | Uberaba | 27999 | 19.0 | 22.9 | 26.4 | 24.4 | 0 | 1.3 |
| | Uberlândia | 43566 | 18.8 | 22.3 | 25.8 | 24.1 | 3.4 | -0.1 |
| | Uruguaiana | 12981 | 10.0 | 20.6 | 28.0 | 25.4 | 6.9 | 1.3 |
| | Varginha | 10080 | 15.5 | 20.4 | 23.9 | 22.3 | -3.7 | 1.4 |
| | Vitória | 123055 | 20.0 | 23.5 | 26.6 | 25.3 | 5.2 | 0 |
| | Vitória da Conquista | 25399 | 17.4 | 21.3 | 24.1 | 22.6 | 6.1 | -0.9 |

| | | | Temperature | | | Temperature Association with Mortality | | |
|---|---|---|---|---|---|---|---|---|
| Country | City | Total Number of Deaths | 5th Percentile Temperature | Median Temperature | 95th Percentile Temperature | Minimum Mortality Temperature $^{0}$C | Excess Death Fraction All Cold (%) | Excess Death Fraction All Heat (%) |
| | Vitória de Santo Antão | 12508 | 22.0 | 24.3 | 26.1 | 24.5 | -5.8 | 2.4 |
| | Volta Redonda | 55448 | 16.1 | 21.2 | 25.7 | 23.3 | 5.8 | 1.2 |
| **Chile** | Antofagasta | 21398 | 9.0 | 13.7 | 17.2 | 17.9 | 0.8 | -0.1 |
| | Arica | 13061 | 14.1 | 17.2 | 20.8 | 21.4 | 10.8 | -0.1 |
| | Calama | 7436 | 8.0 | 13.4 | 16.7 | 17.6 | 23.2 | 0 |
| | Chillán | 14574 | 6.3 | 13.1 | 21.9 | 19.7 | 8 | -0.6 |
| | Concepción | 62576 | 7.2 | 12.6 | 19.5 | 21.1 | 28.5 | -0.2 |
| | Copiapó | 8457 | 11.5 | 17.5 | 21.1 | 18.8 | 9.1 | -1 |
| | Curicó | 9254 | 6.0 | 13.7 | 22.3 | 23.4 | 45.8 | -0.1 |
| | Iquique | 14099 | 13.6 | 16.9 | 21.1 | 21.9 | 21.2 | -0.1 |
| | La Serena-Coquimbo | 23622 | 11.5 | 16.6 | 20.4 | 21.2 | 24.3 | -0.1 |
| | Los Ángeles | 12309 | 5.9 | 12.6 | 21.5 | 19.5 | 20.5 | 0.5 |
| | Osorno | 13008 | 4.4 | 10.5 | 17.7 | 20.1 | 19.2 | -0.1 |
| | Puerto Montt | 13059 | 5.0 | 10.1 | 16.0 | 18.1 | 22.7 | 0.1 |
| | Punta Arenas | 9676 | 0.6 | 4.9 | 9.2 | 10.8 | -9 | 0.1 |
| | Quillota | 12020 | 8.9 | 15.0 | 20.5 | 18.4 | 2.3 | 2.3 |
| | Rancagua | 20033 | 3.3 | 12.2 | 20.7 | 18.9 | 8.8 | -0.5 |
| | San Antonio | 9559 | 8.5 | 13.7 | 18.5 | 19.6 | 13.9 | -0.1 |
| | Santiago de Chile | 387651 | 5.8 | 14.5 | 22.1 | 23.5 | 15.2 | 0 |
| | Talca | 16898 | 6.7 | 14.2 | 23.1 | 20.9 | 10.1 | -1.1 |

| | | | Temperature | | | Temperature Association with Mortality | | |
|---|---|---|---|---|---|---|---|---|
| Country | City | Total Number of Deaths | 5th Percentile Temperature | Median Temperature | 95th Percentile Temperature | Minimum Mortality Temperature $^{0}C$ | Excess Death Fraction All Cold (%) | Excess Death Fraction All Heat (%) |
| | Temuco | 21901 | 4.9 | 11.1 | 18.8 | 17.3 | 13.4 | 0.8 |
| | Valdivia | 11453 | 5.1 | 10.7 | 17.7 | 20.2 | 18.9 | -0.1 |
| | Valparaíso-Viña del Mar | 74162 | 9.1 | 14.0 | 18.7 | 19.6 | 14.5 | 0 |
| **Costa Rica** | San José | 64117 | 17.8 | 19.4 | 20.7 | 19.8 | 2.5 | 1.2 |
| **El Salvador** | San Miguel | 6601 | 24.6 | 27.1 | 30.1 | 26.6 | 0.6 | 3.2 |
| | San Salvador | 46465 | 21.9 | 23.8 | 25.8 | 26.5 | 36 | -0.3 |
| | Santa Ana | 8751 | 20.7 | 23.2 | 25.6 | 23.8 | 0.8 | -1.4 |
| **Guatemala** | Ciudad de Guatemala | 127016 | 16.3 | 18.8 | 20.8 | 20.1 | 1.7 | -0.1 |
| | Escuintla | 7720 | 22.1 | 23.7 | 25.7 | 24.3 | 7.6 | 1.2 |
| | Quetzaltenango | 10688 | 12.8 | 14.5 | 15.7 | 16.5 | 3.5 | 0 |
| **México** | Acapulco (Acapulco de Juárez) (ZM de Acapulco) | 51823 | 24.0 | 25.8 | 28.0 | 25.1 | 0.8 | 3.7 |
| | Acayucan (ZM de Acayucan) | 6687 | 19.8 | 25.2 | 29.6 | 25.5 | 2.9 | 5.7 |
| | Aguascalientes (ZM de Aguascalientes) | 43701 | 12.3 | 18.4 | 23.6 | 20.4 | 9.5 | 1.4 |
| | Campeche (San Francisco de Campeche) | 14234 | 22.1 | 27.1 | 30.7 | 27.3 | 6.3 | -0.6 |
| | Cancún (ZM de Cancún) | 21975 | 22.3 | 26.3 | 28.5 | 26.4 | 4.1 | -1.7 |
| | Celaya (ZM de Celaya) | 41384 | 13.4 | 18.6 | 23.4 | 20.6 | 10.1 | 0.9 |
| | Chetumal (Othón P. Blanco) | 10943 | 22.2 | 26.1 | 28.7 | 25.6 | -2.2 | 3.6 |
| | Chihuahua (ZM de Chihuahua) | 57734 | 7.4 | 19.6 | 28.4 | 22.9 | 6.4 | 1.2 |
| | Chilpancingo (Chilpancingo de los Bravo) | 9874 | 17.5 | 19.6 | 22.2 | 23 | 15.2 | -0.1 |

| | | | Temperature | | | Temperature Association with Mortality | | |
|---|---|---|---|---|---|---|---|---|
| **Country** | **City** | **Total Number of Deaths** | **5th Percentile Temperature** | **Median Temperature** | **95th Percentile Temperature** | **Minimum Mortality Temperature $^0$C** | **Excess Death Fraction All Cold (%)** | **Excess Death Fraction All Heat (%)** |
| | Ciudad Acuña | 6412 | 8.5 | 22.9 | 32.3 | 26.2 | 16.2 | -1.1 |
| | Ciudad Juárez (Juárez) (ZM de Juárez) | 82978 | 4.5 | 19.2 | 30.9 | 25 | 6.3 | 1.9 |
| | Ciudad Obregón (Cajeme) | 24693 | 15.8 | 25.3 | 33.0 | 28.3 | 6.7 | 1.7 |
| | Ciudad Valles | 9540 | 15.9 | 24.1 | 28.5 | 24.8 | -0.5 | 1.9 |
| | Ciudad Victoria | 16362 | 12.9 | 22.5 | 27.8 | 23.9 | 10.4 | -0.1 |
| | Ciudad de México [Mexico City] (ZM del Valle de México) | 1136469 | 11.0 | 15.2 | 18.8 | 17.4 | 5.7 | 0 |
| | Ciudad del Carmen | 10524 | 22.4 | 26.6 | 30.1 | 25.9 | -0.4 | 0.1 |
| | Coatzacoalcos (ZM de Coatzacoalcos) | 19138 | 21.1 | 25.6 | 29.3 | 24.7 | 4.6 | -0.3 |
| | Colima (ZM Colima-Villa de Álvarez) | 19304 | 20.4 | 23.1 | 25.8 | 23.9 | 3.6 | 1.8 |
| | Cuauhtémoc | 10374 | 5.1 | 15.2 | 22.9 | 18.8 | 2.1 | 1 |
| | Cuautla (ZM de Cuautla) | 24402 | 17.6 | 20.7 | 25.2 | 22.6 | 7 | 2.5 |
| | Cuernavaca (ZM de Cuernavaca) | 53405 | 16.3 | 19.7 | 23.6 | 21.1 | 4.2 | 1.6 |
| | Culiacán (Culiacán Rosales) | 42445 | 18.4 | 25.2 | 29.9 | 25.6 | 3.5 | 3.5 |
| | Córdoba (ZM de Córdoba) | 21392 | 14.2 | 19.8 | 22.6 | 20.4 | 3.8 | 1.6 |
| | Delicias | 9428 | 9.3 | 22.0 | 30.8 | 24.9 | 6.7 | 3.3 |
| | Durango (Victoria de Durango) | 31587 | 9.7 | 17.4 | 23.2 | 20.7 | 6.8 | -0.2 |
| | Ensenada | 24874 | 9.8 | 16.6 | 24.9 | 22.4 | 4.2 | 0.1 |
| | Fresnillo | 11899 | 9.8 | 16.8 | 22.0 | 18.9 | 7.9 | 1.4 |
| | Guadalajara* (ZM de Guadalajara) | 237261 | 15.7 | 20.1 | 24.7 | 22.8 | 6.5 | 0.4 |

| | | | Temperature | | | Temperature Association with Mortality | | |
|---|---|---|---|---|---|---|---|---|
| **Country** | **City** | **Total Number of Deaths** | **5th Percentile Temperature** | **Median Temperature** | **95th Percentile Temperature** | **Minimum Mortality Temperature $^{0}$C** | **Excess Death Fraction All Cold (%)** | **Excess Death Fraction All Heat (%)** |
| | Guanajuato | 8520 | 12.1 | 17.5 | 22.3 | 20 | 13.6 | 0.8 |
| | Guaymas (Heroica Guaymas) (ZM de Guaymas) | 13274 | 16.0 | 24.4 | 32.3 | 29.7 | 4 | 2.1 |
| | Hermosillo | 40804 | 13.8 | 24.8 | 33.8 | 30.9 | 14.9 | 1.4 |
| | Hidalgo del Parral | 8168 | 8.4 | 18.6 | 26.0 | 21.5 | 13.1 | -0.8 |
| | Iguala (Iguala de la Independencia) | 9537 | 22.5 | 25.4 | 30.3 | 24.2 | 0.6 | 0.1 |
| | Irapuato | 26970 | 14.3 | 19.1 | 24.4 | 21.4 | 6 | 1 |
| | La Paz | 13197 | 16.9 | 23.8 | 29.8 | 26.5 | 1.8 | 3.2 |
| | La Piedad (La Piedad de Cabadas) (ZM de La Piedad-Pénjamo) | 15474 | 14.4 | 18.8 | 24.1 | 21.2 | -2.7 | 0.6 |
| | León (León de los Aldama) (ZM de León) | 73368 | 13.2 | 18.6 | 23.9 | 20.9 | 8.5 | 0 |
| | Los Mochis (Ahome) | 21853 | 17.4 | 25.2 | 31.5 | 27.7 | 10.5 | 0.7 |
| | Manzanillo | 7900 | 22.4 | 25.1 | 27.2 | 25.5 | -4.4 | 2.9 |
| | Matamoros (Heroica Matamoros) (ZM de Matamoros) | 23848 | 13.8 | 24.8 | 29.6 | 25.4 | 2.6 | 0.9 |
| | Mazatlán | 24221 | 19.5 | 24.7 | 28.4 | 25.3 | 6.4 | 4 |
| | Mexicali (ZM de Mexicali) | 52603 | 12.4 | 24.3 | 36.6 | 33.5 | 7.7 | 2 |
| | Minatitlán (ZM de Minatitlán) | 21364 | 20.8 | 25.5 | 29.5 | 23.9 | 2.7 | 4.4 |
| | Monclova (ZM Monclova-Frontera) | 18667 | 9.2 | 22.6 | 30.4 | 24.9 | 15.5 | 3.2 |
| | Monterrey (ZM de Monterrey) | 213821 | 10.5 | 21.9 | 28.6 | 23.7 | 7.3 | 1.9 |
| | Morelia (ZM de Morelia) | 42626 | 13.0 | 16.7 | 20.6 | 18.4 | 2.6 | 0.7 |
| | Mérida (ZM de Mérida) | 57995 | 21.4 | 26.9 | 30.4 | 25.6 | 0.5 | 2.5 |

| | | | Temperature | | | Temperature Association with Mortality | | |
|---|---|---|---|---|---|---|---|---|
| Country | City | Total Number of Deaths | 5th Percentile Temperature | Median Temperature | 95th Percentile Temperature | Minimum Mortality Temperature $^{0}$C | Excess Death Fraction All Cold (%) | Excess Death Fraction All Heat (%) |
| | Navojoa | 10216 | 16.4 | 25.5 | 32.3 | 27.8 | 6.5 | -0.3 |
| | Nogales (Heroica Nogales) | 9549 | 5.5 | 18.2 | 28.3 | 23.1 | -5.7 | 0.3 |
| | Nuevo Laredo (ZM de Nuevo Laredo) | 20722 | 10.8 | 25.2 | 32.7 | 26.6 | 7.4 | 4.5 |
| | Oaxaca (Oaxaca de Juárez) (ZM de Oaxaca) | 30204 | 13.9 | 17.9 | 21.5 | 19.7 | 5.3 | 0.2 |
| | Ocotlán (ZM de Ocotlán) | 8589 | 16.0 | 20.2 | 24.8 | 22.7 | 7.6 | 3 |
| | Orizaba (ZM de Orizaba) | 29656 | 11.8 | 17.6 | 20.1 | 19.5 | 11 | -0.1 |
| | Pachuca (Pachuca de Soto) (ZM de Pachuca) | 25139 | 8.8 | 13.6 | 17.1 | 18.4 | 3.9 | 0 |
| | Piedras Negras (ZM Piedras Negras) | 10140 | 9.3 | 23.9 | 33.1 | 26.9 | 6.8 | 0.5 |
| | Playa del Carmen | 4270 | 22.0 | 26.2 | 28.5 | 25.8 | 0.5 | 2.5 |
| | Poza Rica de Hidalgo (ZM de Poza Rica) | 36768 | 16.7 | 24.1 | 27.6 | 24.3 | 1.9 | 2.4 |
| | Puebla (Heróica Puebla de Zaragoza) (ZM Puebla) | 154580 | 12.5 | 16.1 | 19.3 | 20.5 | 10.1 | 0 |
| | Puerto Vallarta (ZM de Puerto Vallarta) | 15401 | 20.1 | 24.1 | 26.7 | 23.9 | 1.1 | 7.8 |
| | Querétaro (Santiago de Querétaro) (ZM de Querétaro) | 49050 | 12.3 | 17.8 | 22.4 | 19.7 | 5.5 | -0.4 |
| | Reynosa (ZM de Reynosa-Río Bravo) | 34077 | 12.7 | 25.2 | 30.8 | 25.9 | 5.7 | -0.5 |
| | Rioverde (Río Verde) (ZM de Río Verde-Ciudad Fernández) | 9291 | 12.1 | 20.5 | 25.0 | 21.7 | 11.2 | 2.3 |
| | Salamanca | 14250 | 14.5 | 19.4 | 24.6 | 21.5 | 6.6 | 0.7 |
| | Saltillo (ZM de Saltillo) | 39429 | 8.4 | 17.7 | 22.7 | 19.5 | 13.4 | -0.3 |
| | San Cristóbal de las Casas | 7780 | 12.8 | 15.7 | 17.9 | 18.8 | 18.5 | 0 |

| | | | Temperature | | | Temperature Association with Mortality | | |
|---|---|---|---|---|---|---|---|---|
| Country | City | Total Number of Deaths | 5th Percentile Temperature | Median Temperature | 95th Percentile Temperature | Minimum Mortality Temperature $^0$C | Excess Death Fraction All Cold (%) | Excess Death Fraction All Heat (%) |
| | San Francisco del Rincón (ZM de San Francisco del Rincón) | 8593 | 13.6 | 18.8 | 24.1 | 21 | 5.1 | -0.1 |
| | San Juan Bautista Tuxtepec | 7914 | 19.8 | 25.4 | 29.9 | 26.3 | 2.5 | 0.1 |
| | San Juan del Río | 10608 | 11.4 | 16.8 | 21.3 | 18.5 | 13.2 | 0.3 |
| | San Luis Potosí (ZM de San Luis Potosí-Soledad de Graciano Sánchez) | 50439 | 9.9 | 17.1 | 21.7 | 18.8 | 10.1 | -0.6 |
| | San Luis Río Colorado | 9784 | 11.9 | 24.0 | 35.6 | 32.3 | 5.3 | 3.8 |
| | Santo Domingo Tehuantepec (ZM de Tehuantepec) | 9298 | 22.7 | 26.5 | 29.3 | 25.8 | -4.5 | 7.4 |
| | Tampico (ZM de Tampico) | 49930 | 17.7 | 25.2 | 28.5 | 25.1 | 4.4 | 2.1 |
| | Tapachula (Tapachula de Córdova y Ordóñez) | 19418 | 22.9 | 24.5 | 26.4 | 24.9 | -0.6 | -1.4 |
| | Tecomán (ZM Tecomán) | 8960 | 23.1 | 25.6 | 27.8 | 25.8 | 0.1 | 3.3 |
| | Tehuacán (ZM de Tehuacán) | 14686 | 12.2 | 17.7 | 21.3 | 19.4 | 9.5 | -0.9 |
| | Tepic (ZM de Tepic) | 21261 | 16.3 | 21.0 | 23.5 | 21.6 | 4.8 | 1.9 |
| | Teziutlán (ZM de Teziutlán) | 6796 | 9.2 | 14.6 | 17.5 | 18.8 | 31.2 | -0.2 |
| | Tianguistenco (ZM de Tianguistenco) | 9026 | 8.9 | 12.6 | 15.2 | 16.5 | -1.7 | -0.1 |
| | Tijuana* (ZM e Tijuana) | 84415 | 9.9 | 16.1 | 23.5 | 21.9 | 9.3 | 0.2 |
| | Tlaxcala (Tlaxcala de Xicohténcatl) (ZM de Tlaxcala-Apizaco) | 24472 | 10.2 | 14.4 | 17.7 | 19 | 15.9 | 0 |
| | Toluca (Toluca de Lerdo) (ZM de Toluca) | 94146 | 9.5 | 13.2 | 16.2 | 17.5 | -2.3 | 0 |
| | Torreón (ZM de la Laguna) | 69230 | 12.2 | 23.5 | 29.9 | 25 | 10.1 | 3.1 |

| | | | Temperature | | | Temperature Association with Mortality | | |
|---|---|---|---|---|---|---|---|---|
| **Country** | **City** | **Total Number of Deaths** | **5th Percentile Temperature** | **Median Temperature** | **95th Percentile Temperature** | **Minimum Mortality Temperature $^{0}$C** | **Excess Death Fraction All Cold (%)** | **Excess Death Fraction All Heat (%)** |
| | Tula de Allende (ZM de Tula) | 13777 | 10.6 | 15.7 | 19.8 | 21.2 | 21.5 | 0 |
| | Tulancingo (Tulancingo de Bravo) (ZM de Tulancingo) | 12001 | 8.3 | 13.8 | 16.8 | 18.1 | -19.1 | 0 |
| | Tuxtla Gutiérrez (ZM Tuxtla Gutiérrez) | 32312 | 18.1 | 22.5 | 26.2 | 23.4 | 1.1 | 0.2 |
| | Uriangato (ZM de Moroleón-Uriangato) | 6582 | 14.0 | 18.1 | 23.0 | 21.2 | 3.1 | -1.4 |
| | Uruapan (Uruapan del Progreso) | 17063 | 14.7 | 17.9 | 20.9 | 19.2 | 5.3 | 1.9 |
| | Veracruz (ZM de Veracruz) | 53342 | 20.3 | 25.6 | 29.0 | 25.1 | -1 | 1.8 |
| | Villahermosa (ZM de Villahermosa) | 37912 | 21.4 | 26.2 | 31.1 | 25.1 | -0.8 | 5.7 |
| | Xalapa-Enríquez (Jalapa) (ZM de Xalapa) | 38085 | 13.3 | 19.4 | 22.2 | 23.7 | 24.1 | -0.1 |
| | Zacatecas (ZM de Zacatecas-Guadalupe) | 13948 | 9.7 | 16.2 | 20.9 | 18.2 | 5.6 | 0.8 |
| | Zamora de Hidalgo (ZM de Zamora-Jacona) | 14641 | 14.9 | 18.9 | 23.9 | 20.9 | -4.7 | 2.2 |
| **Panamá** | Colon | 4737 | 24.8 | 25.7 | 26.7 | 27.3 | -11 | 0.2 |
| | David | 4267 | 23.5 | 24.9 | 27.4 | 25.5 | 9.9 | 1.1 |
| | Panama City | 32524 | 24.7 | 25.8 | 27.1 | 25.4 | -0.4 | -0.7 |
| **Perú** | Arequipa | 29148 | 11.4 | 13.6 | 16.1 | 17.2 | 7.1 | 0 |
| | Ayacucho | 4833 | 9.4 | 11.6 | 13.4 | 14 | 26.5 | -0.1 |
| | Cajamarca | 4120 | 10.2 | 11.6 | 13.0 | 13.6 | 64.1 | -1.3 |
| | Chiclayo | 22919 | 18.0 | 21.1 | 25.7 | 23.8 | 3.1 | 0.2 |
| | Chimbote | 8851 | 16.2 | 19.2 | 24.0 | 22.5 | 20.4 | -0.8 |
| | Chincha Alta | 6544 | 17.0 | 19.5 | 22.9 | 21.3 | 2.1 | 3.1 |

| | | | Temperature | | | Temperature Association with Mortality | | |
|---|---|---|---|---|---|---|---|---|
| Country | City | Total Number of Deaths | 5th Percentile Temperature | Median Temperature | 95th Percentile Temperature | Minimum Mortality Temperature $^{0}C$ | Excess Death Fraction All Cold (%) | Excess Death Fraction All Heat (%) |
| | Cusco (Cuzco) | 7376 | 6.1 | 8.0 | 9.7 | 10.6 | -7.3 | 0.1 |
| | Huancayo | 16284 | 6.9 | 8.7 | 10.2 | 10.8 | -1.9 | -0.1 |
| | Huaraz | 2971 | 6.4 | 8.0 | 9.5 | 10.1 | 77.6 | -1 |
| | Huánuco | 6274 | 12.5 | 14.1 | 15.4 | 16 | -0.9 | -0.3 |
| | Ica | 10458 | 17.9 | 21.2 | 24.3 | 22.6 | 1.2 | -0.1 |
| | Iquitos | 4663 | 23.9 | 25.5 | 27.4 | 24.8 | 1.7 | 16.8 |
| | Juliaca | 9400 | 5.6 | 8.4 | 11.1 | 12.2 | 1.4 | 0 |
| | Lima | 261923 | 15.9 | 18.6 | 22.2 | 21.1 | 6.9 | 0.5 |
| | Pisco (incl. San Clemente) | 3862 | 16.4 | 19.5 | 23.9 | 22.4 | 15.6 | -0.9 |
| | Piura | 15607 | 20.6 | 23.9 | 27.6 | 25.7 | 6.8 | 1.1 |
| | Pucallpa | 9719 | 23.7 | 25.7 | 27.9 | 24.8 | 0.2 | 11.3 |
| | Puno | 4875 | 5.2 | 8.0 | 10.5 | 11.6 | 30.7 | -0.4 |
| | Sullana | 8959 | 20.8 | 24.0 | 27.3 | 25.5 | 3.3 | 0.7 |
| | Tacna | 7822 | 13.8 | 17.4 | 20.8 | 19.3 | -9.7 | 0.8 |
| | Tarapoto | 3535 | 22.5 | 24.1 | 26.0 | 24.3 | -6.2 | 0.1 |
| | Trujillo | 30797 | 16.9 | 19.4 | 22.7 | 20.8 | -1.8 | 1.6 |
| | Tumbes | 2518 | 22.7 | 24.8 | 26.5 | 25.3 | -2.8 | -0.7 |
